# Evaluation of inter-observer agreement for periportal fibrosis related to *Schistosoma mansoni*: A population-based study of 3302 individuals using point-of-care ultrasound images and video in the SchistoTrack cohort

**DOI:** 10.64898/2026.11.02.26367045

**Authors:** Eloise Ockenden, Chris Ho, Victor Anguajibi, Simon Mpooya, Tymothy Mugume, Benjamin Ntegeka, Robert Ntakimanye, Ivan Tumukunde, Betty Nabatte, Narcis B. Kabatereine, Goylette F. Chami

**Author notes:** These authors contributed equally to this work (Eloise Ockenden, Chris Ho).

## Abstract

**Background:** Schistosomiasis-related periportal fibrosis (PPF) diagnoses rely on point-of-care ultrasound imaging. Yet, the extent of agreement between sonographers, limitations of existing World Health Organization (WHO) protocols, and the applicability of ultrasound video remain poorly understood.

**Methods:** In 2024, six sonographers performed ultrasound scans for 3302 participants aged 5–92 years within SchistoTrack, a community-based cohort in rural Uganda. We adapted and standardised the WHO Niamey protocol for the collection of curated ultrasound images of liver fibrosis patterns related to *Schistosoma mansoni* (ranging A–F) and liver ultrasound video sweeps. PPF was defined as patterns C–F. Images and videos were read for patterns of fibrosis by second sonographers who were blinded to field diagnoses. Second sonographers recorded their clinical confidence, the probe position and any ultrasound interference. Outcomes considered were simple agreement for individual patterns and agreement on whether any PPF pattern was present, using Cohen’s ***κ***. The influence of participant characteristics and image quality factors was evaluated against agreement outcomes.

**Results:** Abnormal liver fibrosis patterns (B–F) were identified in 32.8% (1082/3302) of participants, and among these 2074 images were read by a second sonographer from 1040 participants. The percentage agreement was 73.0% (1515/2074) for broad pattern categories. Severe fibrosis patterns had the least agreement; E and F patterns had ***κ*** = 0.430 and 0.149, respectively. The D pattern had the most agreement (***κ*** = 0.717). 94.0% of second sonographer readings were within one step of severity of the field diagnosis. Participant characteristics were not consistently associated with disagreement, though agreement clustered by the field sonographer. Clinical confidence, breathing/movement, fat interference and probe position were positively associated with disagreement. There was no agreement among the 1054 videos that were read by a second sonographer: ***κ*** = 0.001 for PPF agreement and ranged 0–0.062 for individual patterns.

**Conclusion:** We have presented the largest diagnostic study of inter-sonographer agreement for schistosomal liver fibrosis patterns. Disagreement between adjacent patterns of liver fibrosis may indicate that the WHO Niamey protocol should be updated to capture a continuous progression of liver fibrosis rather than distinct patterns. The greatest disagreement was among severe patterns, which supports routine second reading to ensure life-threatening cases are prioritised and appropriately triaged.

## Background

Schistosomiasis is a water-borne parasitic disease, which is one of the most prevalent neglected tropical diseases (NTDs) worldwide, estimated to affect over 250 million people–most of whom are in sub-Saharan Africa [1]. Among the different species of *Schistosoma* that cause human infection, *S. mansoni* is one of the most prevalent in sub-Saharan Africa [2]. If left untreated, chronic *S. mansoni* infection is associated with complications such as periportal fibrosis (PPF) [3], characterised by pathological scarring around the portal vein of the liver. PPF may result in portal hypertension and its associated sequelae, which include ascites and portosystemic collaterals [4]. PPF can be detected using point-of-care ultrasound imaging (POCUS) [5].

The WHO Niamey protocol, introduced in 1996, describes a semi-standardised system to observe PPF using POCUS [6]. The system mainly focuses on the use of image patterns representing presentations of PPF [7]. The patterns (A–F) span diffuse and focal presentations of liver fibrosis, where the A pattern represents a healthy liver and the F pattern represents fibrosis extending to the liver capsule. B patterns are diffuse patterns that are not necessarily associated with schistosomiasis. C patterns represent diffuse fibrosis in the second order portal branches, and the D pattern represents fibrosis along the main portal vein. The E pattern is visualised as patches of fibrosis representing blocked vessels wit no lumen. Whilst the WHO Niamey protocol has been generally well accepted since its publication [8], it lacks clear implementation guidance and requires specific expertise to implement. The protocol lacks instructions on how to acquire data, including matching specific probe positions to where liver fibrosis patterns are likely to be found. The protocol also lacks guidance on how to validate results, which may lead to low reproducibility of studies using this method of liver fibrosis assessment. The need for clear guidance is even greater in low- and middle-income country (LMIC) settings, where prior literature suggests difficulties including lack of access to training, equipment, and machine maintenance capability [9–11].

Understanding the inter-observer variability of a diagnostic tool is important to assess potential measurement error or misdiagnosis in clinical and research settings. Understanding agreement for the WHO Niamey protocol image patterns present different challenges exacerbated in low-income settings. Inter-observer agreement has been quantified throughout development of staging systems for PPF diagnosis [7]. Before the development of the WHO Niamey protocol, inter-observer agreement studies were carried out on measurement-based systems focused on enlargement of the segmental portal branches, which preceded the WHO Niamey protocol. For example, studies using the Managil protocol [12] found measurement-based agreement to be fair to moderate (Cohen’s *κ* ranging 0.29–0.59) [13–15]. By contrast, studies looking at liver fibrosis patterns across an image using the WHO Niamey protocol [6] generally found agreement to be higher than for the measurement-based systems (Cohen’s *κ* ranged 0.46–0.89) [16–19]. However, the methods used to measure agreement were highly heterogeneous. Some studies compared observers that both scanned participants [16, 17]. Other studies compared patterns assigned at dynamic examinations to assignments to static images [16, 19]. This heterogeneity makes it difficult to definitively establish how reproducible or reliable the WHO Niamey protocol image patterns are for diagnosis of PPF. Additionally, the patterns have not been validated in clinical settings since they were developed for research. Limitations of existing studies include the failure to acknowledge variation in image quality, the presence of potential artifacts, and the influence of participant characteristics or potential comorbidities on interobserver agreement. In other fields, image quality has been shown to affect agreement for endoscopy [20] and digital mammography [21]. Patient-level factors such as age and possible comorbidities also have been shown to affect ultrasound image quality [22, 23]. Critically, for clinical settings, clinical confidence in the diagnosis provided must be assessed to understand how useful the diagnosis assigned will be for case management or patient follow-up. To our knowledge, no studies on schistosomiasis have assessed clinical confidence. Comprehensive, large-scale standardised inter-observer agreement studies are needed to inform future updates of the WHO Niamey protocol.

To our knowledge, we have conducted the largest, most comprehensive study of inter-observer agreement for schistosomal liver fibrosis associated with *S. mansoni* using ultrasound images and video. We examined 3302 individuals aged five years and older within the rural, community-based SchistoTrack cohort. We provide a detailed, repeatable protocol that improves upon the WHO Niamey protocol to enable the assessment of inter-observer agreement of schistosomal liver fibrosis patterns. The main aim of this study was to provide inter-observer agreement metrics for schistosomal liver fibrosis patterns assessed through ultrasound image and video while describing any image, participant-level, or observer factors that may influence disagreement.

## Methods

### Study context and participant sampling

This study was conducted within the 2024 annual time point of SchistoTrack, a longitudinal cohort study which assesses morbidity, transmission, and treatment of schistosomiasis in Uganda [24]. The SchistoTrack cohort is in 52 villages across three rural districts of Uganda: Pakwach, Buliisa and Mayuge, where the prevalence of *S. mansoni* exceeds 35%, and is up to 50% in Pakwach [25]. 30 households from each village were randomly sampled, with oversampling to account for non-participation. From each household, one participant aged *≥* 18 years old and one participant aged *<* 18 years old were selected for clinical data collection by the household head or lead wife. Selected participants attended ultrasound imaging data collection as part of wider clinical data collection annually. Ultrasound imaging data collection was done on a bed behind privacy curtains, inside a building such as a school. Participants were told in advance to arrive at the ultrasound station having fasted for at least two hours.

### Sonography team and equipment

The POCUS protocol was designed jointly by the study team and the Ugandan sonography team. The sonography team comprised six sonographers currently working in clinical practice and with 5–20 years of experience using the WHO Niamey protocol.

Ultrasound scans were performed using portable Philips C5-2 curved array transducers [26] connected via universal serial bus (USB) to Android (version 9 Pie) tablets running the Lumify [27] application. Lumify integrates the transducers with the Android tablets, permitting ultrasound videos and images to be saved onto the tablets’ Secure Digital (SD) cards, and facilitates annotating the videos/images with text labels. Abdomen View was selected as the scan option in Lumify. All videos and images were saved in Digital Imaging and COmmunications in Medicine (DICOM) [28] format. Electronic surveys were performed on Android (version 9 Pie) tablets using the Open Data Kit (ODK) Collect [29] survey-based data gathering application. Data collected via the ODK survey were streamed to a server for storage and later analysis. For each participant, one sonographer performed the ultrasound scan, recorded video sweeps, and acquired static curated images. An auxiliary worker who spoke the local language of the study village was present to fill out participant information in ODK.

### Video sweeps

The first part of the ultrasound examination was the acquisition of three five-second-long ultrasound video sweeps capturing views of the liver and surrounding anatomies. The WHO Niamey protocol provides no guidance on ultrasound video. The sweeps were designed to capture all necessary anatomies and views to diagnose periportal fibrosis. Sonographers were instructed to only start the video recordings once a good view of the target anatomy had been found. The sweeps were as follows:

1. *Transverse substernal view.* Starting with the probe just below the sternum in transverse position aligning with the mid-clavicular line, adjust the probe position until there is a clear view of the gall bladder. If the gall bladder is not visible at all, then adjust the probe to where the gall bladder would be located approximately. Begin recording, and perform a sweep moving the probe to the cephalic position, then end the recording.
2. *Right oblique view.* Starting with the probe in the right oblique postion (45°) aiming at the main portal vein, adjust the probe so that the main portal vein has appeared in clear view. Begin recording, and from the main portal vein view, continue in oblique view and turn the probe to transverse to view the first three branches, then end the recording.
3. *Sagittal left parasternal view.* Place the probe in the left sagittal parasternal position (90°) and ensure there is a clear view of the left lobe of the liver – usually in healthy participants, this will be the entire lobe, but may not be the case in participants with lobe enlargement. Begin recording, and sweep the probe until the edge of the left lobe is reached, ensuring that the sweep shows the entire left liver lobe, then end the recording.

### Image acquisition

Following the video sweep recordings, sonographers were instructed to save images of liver fibrosis patterns, and measurements of specified anatomies such as the main portal vein diameter and portal branches. The measurements are beyond the scope of this analysis. Sonographers in our study were instructed to identify any presenting abnormal (non-A) fibrosis patterns during a scan, and capture a still image of a clear view of the pattern. They subsequently labelled the image with B–F, using the Lumify application. Diagnosis of PPF was considered to be assignment of liver patterns C–F. The liver fibrosis patterns to be identified were based on those defined in the WHO Niamey protocol, but were more clearly defined within the SchistoTrack study with clear instructions provided on probe positions and views to find the patterns (Table 1).

**Table 1:**
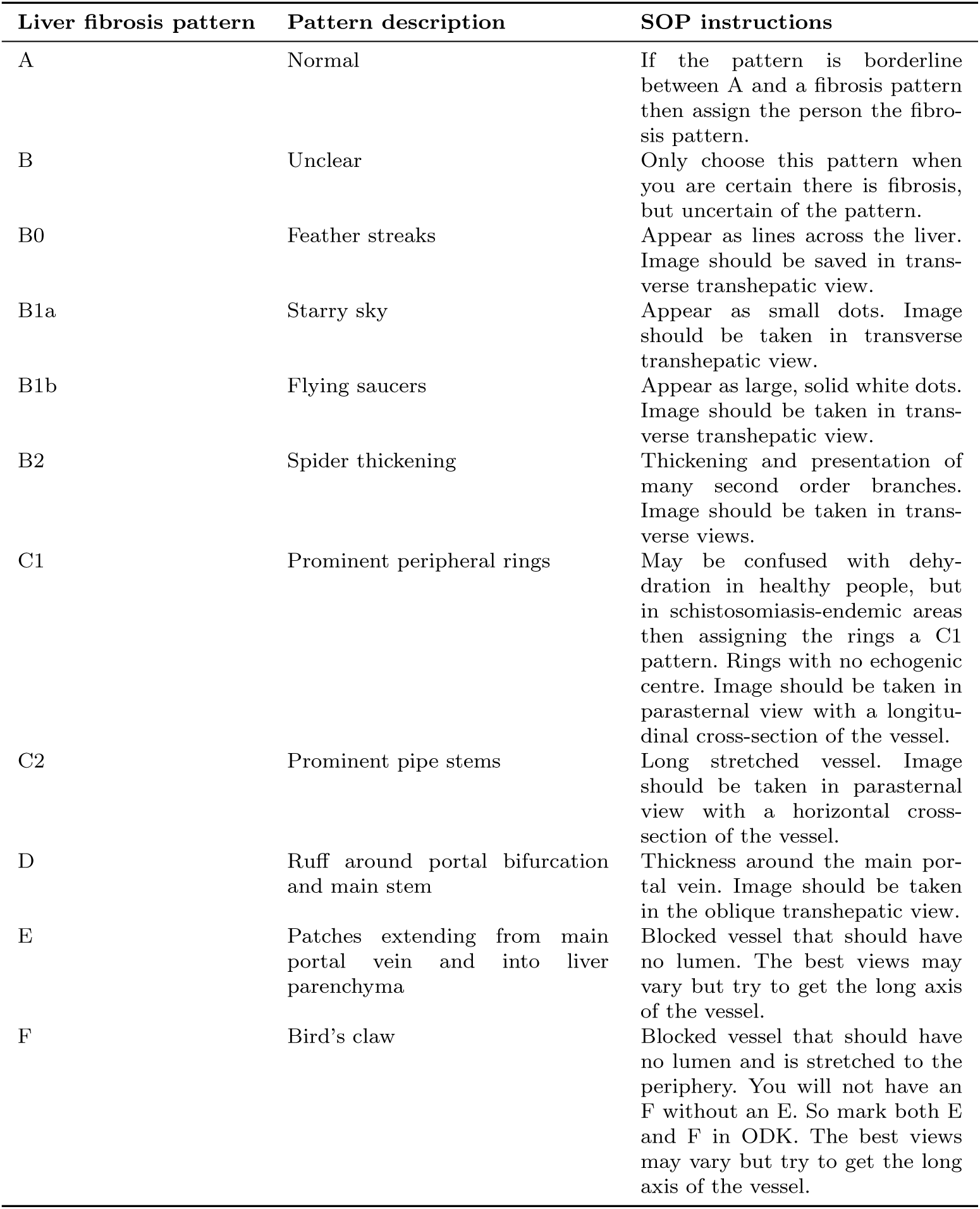
WHO Niamey protocol image patterns. Definitions of each image pattern from the WHO Niamey protocol with SchistoTrack standard operating procedure (SOP) instructions for acquisition.

| **Liver fibrosis pattern** | **Pattern description** | **SOP instructions** |
|---|---|---|
| A | Normal | If the pattern is borderline between A and a fibrosis pattern then assign the person the fibro-sis pattern. |
| B | Unclear | Only choose this pattern when you are certain there is fibrosis, but uncertain of the pattern. |
| B0 | Feather streaks | Appear as lines across the liver. Image should be saved in trans-verse transhepatic view. |
| B1a | Starry sky | Appear as small dots. Image should be taken in transverse transhepatic view. |
| B1b | Flying saucers | Appear as large, solid white dots. Image should be taken in trans-verse transhepatic view. |
| B2 | Spider thickening | Thickening and presentation of many second order branches. Image should be taken in trans-verse views. |
| C1 | Prominent peripheral rings | May be confused with dehy-dration in healthy people, but in schistosomiasis-endemic areas then assigning the rings a C1 pattern. Rings with no echogenic centre. Image should be taken in parasternal view with a longitu-dinal cross-section of the vessel. |
| C2 | Prominent pipe stems | Long stretched vessel. Image should be taken in parasternal view with a horizontal cross-section of the vessel. |
| D | Ruff around portal bifurcation and main stem | Thickness around the main por-tal vein. Image should be taken in the oblique transhepatic view. |
| E | Patches extending from main portal vein and into liver parenchyma | Blocked vessel that should have no lumen. The best views may vary but try to get the long axis of the vessel. |
| F | Bird’s claw | Blocked vessel that should have no lumen and is stretched to the periphery. You will not have an F without an E. So mark both E and F in ODK. The best views may vary but try to get the long axis of the vessel. |

Table 2 summarises the views and expected fibrosis patterns and anatomies appearing in the resultant scans.

**Table 2:**
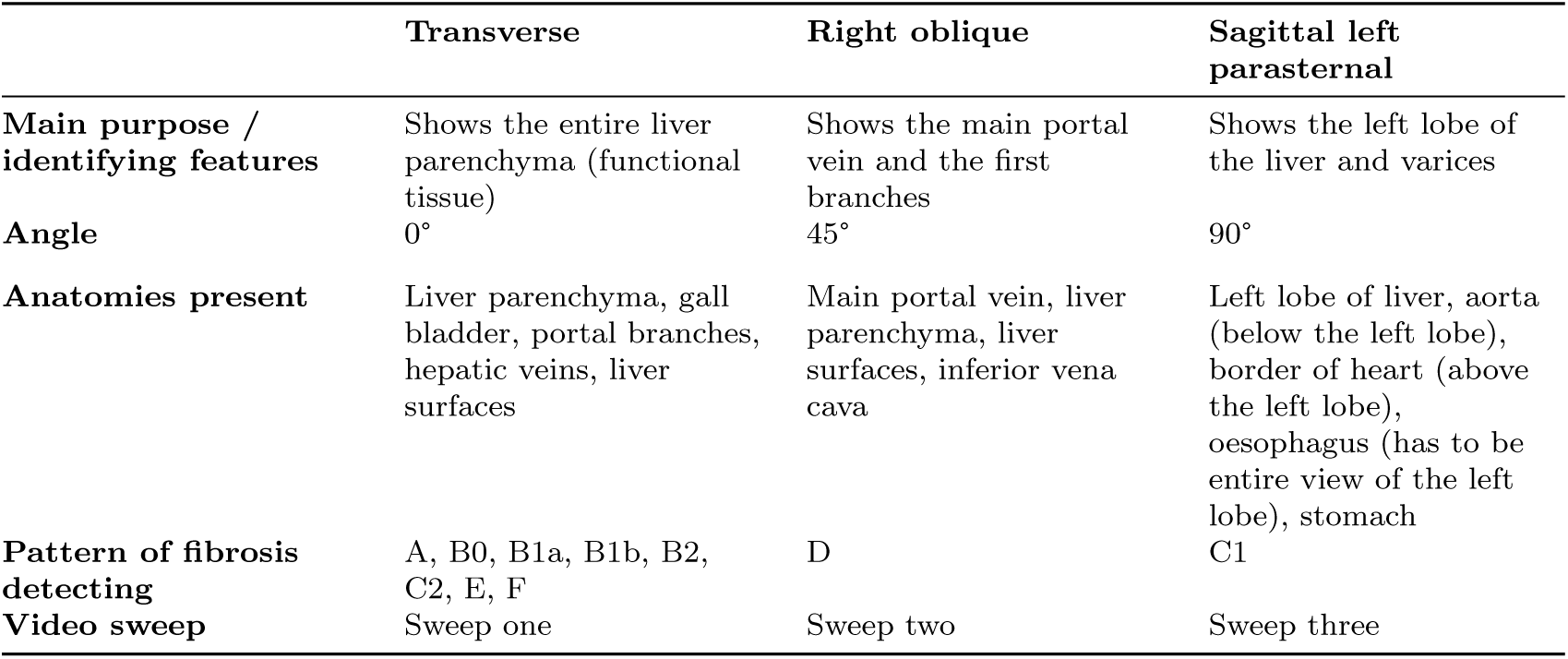
Views of the liver. Summary of views of the liver for PPF diagnosis, with expected anatomies for each view and patterns that can be found in each view.

### Data processing

All ultrasound scans were saved in DICOM format, which requires specialist software to view. The images and videos were therefore converted into the usable Joint Photographic Experts Group (JPEG) and Moving Picture Experts Group-4 (MP4) file formats for second sonographer reading, using the Python programming language (version 3.9.6) and packages pydicom, OpenCV and FFmpeg [30–33].

As previously described, sonographers labelled images of fibrosis liver patterns with the letters B–F. To prepare for assessment of inter-observer agreement on fibrosis grading, we masked the images to hide the liver pattern labels that were outside of the field of view. The motivation of this was to permit blind reading, where the second sonographer would not be influenced by the initial diagnosis. Masking was performed using a contouring image processing algorithm.

### Second sonographer reading of liver fibrosis images and videos

Four sonographers of the six that performed field scanning took part in second sonographer reading. Blind reading of fibrosis images was performed by a randomly selected second sonographer such that no sonographer would be reading any images that they themselves acquired in the field. Participants with abnormal (non-A pattern) liver patterns had their images read by a second sonographer. There were no images of A (normal) liver fibrosis patterns since there was no specific view of this pattern that could be captured. PDFs of images were used with spreadsheets to record patterns fibrosis present in an image. Field sonographers were allowed to assign one liver fibrosis pattern per image to display what they thought was the best represented liver fibrosis pattern in that static image. However, second sonographers who read the images were allowed to assign multiple liver fibrosis patterns to capture all patterns they believed to be represented by the image. Sweep one videos and a subset of sweep two and three videos were converted to MP4 format and played by the sonographers using standard software. The sonographers were asked to record every pattern of fibrosis that appeared in that video in spreadsheets. Due to logistical constraints, sweep one was focused on because it captured the curated views of the most patterns (B0, B1a, B1b, B2, C2, E and F).

For all images and videos, sonographers also reported on interference, clinical confidence, probe position (for images) and how well the sweep protocol was followed (for videos). The interference types that were given and their reasons for inclusion as given in the SOP were as follows. Gas may cause posterior enhancement. Fat may cause excessive brightness in an image. Breathing/movement may cause blurring. Bone can be seen with a posterior shadow. Calcification and dehydration may appear with excessive speckle. Sonographers could also add other types of interference in free text, and add any other comments. The sonographers were asked to rate their clinical confidence for each diagnosis given. Low confidence was defined as “you would not use this video to take clinical actions; this would require another scan”. Medium confidence was defined as “you would use this video to take clinical actions; but more ultrasound information would be helpful.” High confidence was defined as “you would use this video to take clinical actions and it is clear what actions are needed.” Finally, for the images the probe position was recorded or noted as unclear, and for the videos the sonographers recorded whether the sweep procedure was followed and whether the sweep was performed at the appropriate speed to fill the five seconds recommended in the SOP.

### Inter-observer agreement outcomes

We defined several outcome measures based on standard inter-observer agreement metrics, and performed statistical tests to understand the effect of participant characteristics and other biomedical variables on these outcomes. Different outcomes were defined to evaluate agreement on image, video and participant levels.

The outcomes used in this study were specific liver fibrosis pattern agreement, any PPF agreement and numeric differences in liver fibrosis patterns (Table 3). Pattern agreement was assessed at the image level, and was a binary variable equal to one if the field pattern for the image was present in the set of patterns assigned by the second sonographer. There could be multiple patterns present in an image. Consequently, we proposed that the field pattern being present in the set of follow-up diagnoses constituted agreement.

**Table 3:**
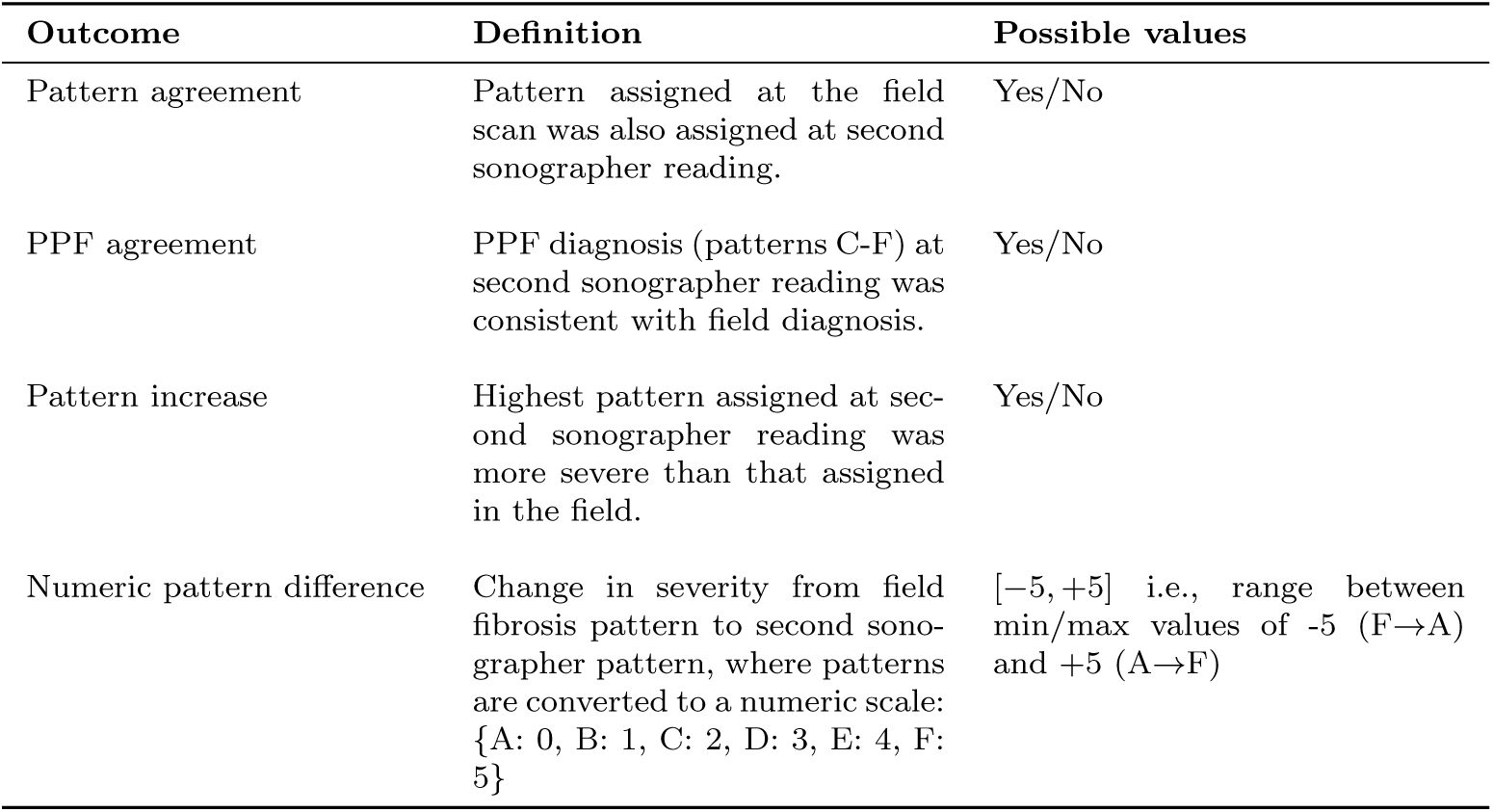
Outcome definitions and ranges of permissible values.

| **Outcome** | **Definition** | **Possible values** |
|---|---|---|
| Pattern agreement | Pattern assigned at the field scan was also assigned at second sonographer reading. | Yes/No |
| PPF agreement | PPF diagnosis (patterns C-F) at second sonographer reading was consistent with field diagnosis. | Yes/No |
| Pattern increase | Highest pattern assigned at sec-ond sonographer reading was more severe than that assigned in the field. | Yes/No |
| Numeric pattern difference | Change in severity from field fibrosis pattern to second sono-grapher pattern, where patterns are converted to a numeric scale: {A: 0, B: 1, C: 2, D: 3, E: 4, F: 5} | [−5, +5] i.e., range between min/max values of -5 (F→A) and +5 (A→F) |

We evaluated individual pattern agreement at two levels of detail to assess whether more discrete patterns would result in higher disagreement. We first used a granular system where B-patterns were separated into B, B0, B1, B2 and C patterns into C1, C2; and secondly used a broader system where each of the individual B and C patterns were grouped into B and C, respectively. The granular patterns represent different extents, severity, or types of liver fibrosis, whereas the broader categories group the patterns of equivalent PPF severity; the aim was to evaluate agreement for both specific patterns and overall potential severity of liver fibrosis. PPF agreement was developed as an outcome to identify cases where disagreement is clinically important, as the treatment for non-schistosomal (B-pattern) fibrosis will be different to schistosomal PPF (patterns C–F) [34]. We assessed PPF agreement at participant level, where a PPF diagnosis was the assignment of a C–F pattern for any image.

The numeric difference in assigned patterns was evaluated between field and second sonographer readings. Here we assumed that the liver fibrosis patterns from A–F were ordered by severity. Given sonographers were allowed to assign multiple followup patterns to a single image, we aggregated the numeric pattern difference values across all pairs by selecting the smallest numeric pattern difference. The motivation for selecting the minimum difference was that a pair of diagnoses may appear to have a large numeric difference if the image happened to also present a more severe pattern, but was the best view of a liver fibrosis pattern of a lower level of severity. When evaluating numeric difference in assigned patterns at the participant level, the highest (most severe) pattern from the field and second sonographer reading were used, as the sonographers would have had the chance to view all presenting patterns from all images of the participant.

### Image and participant-level factors

A combination of image-specific and participant-level factors were considered for their association with agreement outcomes. Image-specific information was gathered from annotations made by second sonographers, as detailed in Section 3. Clinical confidence, probe position and the individual types of interference were recorded.

Participant-level factors included demographic variables (age in years and sex) as well as body mass index (BMI) 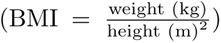. Liver enlargement of left and right lobes of the liver were defined relative to a healthy reference population within SchistoTrack that grouped participants into height deciles, and means and standard deviations (s.d.s) were calculated within-group as described elsewhere [35]. We considered left and right liver lobes to be enlarged/shrunken if they were 2s.d.s above/below the mean for the height interval.

Other participant-level factors included the time of day that the field scan took place, which was grouped into categories morning (8am–12pm) and afternoon (after 12pm–end of the working day). Fasting status (2+ hours) was considered to test if a participant who was not fully fasted affected the second sonographer reading and, in turn, agreement. To understand the effect of hepatic abnormalities identified during the field scan, we considered the presence of any of the following abnormalities or general findings including ascites, pregnancy, cirrhosis-like, fatty-like, or hepatitis-Blike liver; irregular liver surface; rounded liver edge; polycystic kidney disease; liver cysts; situs inversus and portosystemic collaterals (gastro-oesophageal, pancreatico-duodenal, and splenic varices, recanalised paraumbilical veins, and splenorenal shunts). Participants’ infection status with hepatitis B, measured with Abbott rapid diagnostic tests [36], and human immunodeficiency virus (HIV) status measured using STAT PAK [37] and SD Bioline [38] rapid diagnostic tests were recorded. Alcohol use was also considered in four categories. Lifetime non-drinkers were those who had never drunk alcohol; former drinkers were those who had not drunk alcohol in the past year but had drunk alcohol prior to that. Occasional drinkers were those who drunk alcohol less than once a week, and regular drinkers were those who drunk alcohol weekly or daily [39]. The sonographer who performed the field scan was also considered.

### Intra-observer studies

In addition to the inter-observer studies that have been described, we also conducted intra-observer studies on a subset of images and videos to provide a comparison that removes the variability between the readers themselves. A subset of nine participants per sonographer were chosen for intra-observer reading of images, and fourteen participants per sonographer were chosen for intra-observer reading of videos. The chosen participants were selected so that all liver fibrosis patterns were represented in the intra-observer studies.

### Statistical analysis

Analyses were conducted in Python version 3.9.6, using the Pandas, Numpy, Statsmodels, Scikit-Learn, Seaborn, and Matplotlib packages [30, 40–45], and R version 4.2.1. Percentage agreement is reported for each outcome. Cohen’s *κ* is reported, which is a measure of agreement that accounts for chance agreement, and ranges between -1 (complete disagreement) and 1 (complete agreement). Weighted Cohen’s *κ_W_* is reported where data were ordinal, since *κ_W_* assigns higher weights to disagreements that are further apart. Intra-class correlation coefficients (ICCs) for agreement and consistency are reported. ICCs measures the proportion of the variance measurements that is attributable to differences between subjects and as opposed to other factors like diagnostic consistency. ICC for agreement measures absolute agreement for exact categories, whereas for ordinal variables ICC for consistency measures whether the ranking of subjects is the same between raters. Confidence intervals could be calculated for ICC. Tests of significance were used between image-level variables and pattern agreement on an image level and agreement separated out by pattern. For PPF agreement and highest pattern agreement, tests of significance were done between participant level variables and those outcomes. *χ*^2^ tests were used for categorical variables and t-tests were used for continuous variables. Significance was defined as *p <* 0.05.

## Results

### Field PPF prevalence

POCUS scans were performed for 3302 participants from January to February 2024. Female participants comprised 55.03% (1817/3302) of the study population. Ages ranged from 5 to 92 years, with 60.4% (1995/3302) adult participants and 39.6% (1307/3302) children, where adulthood was considered *≥*15 years. Numbers of participants recruited from each of the study districts were: 43.4% (1432/3302) from Pakwach; 33.1% (1094/3302) from Buliisa; and 23.5% (776/3302) from Mayuge.

Table 4 displays the frequency and prevalence of each fibrosis liver pattern among study participants. Field scans identified abnormal liver patterns (B–F) in 32.8% (1082/3302) participants. Periportal fibrosis (PPF) (C-F) was diagnosed in 18.5% (611/3302) participants overall; of the participants with PPF, 82.3% (503/611) were adults and 17.7% (108/611) were children. Male participants constituted 53.5% (327/611) of those with PPF. In the study districts, the prevalence of PPF was 26.4% (378/1432) in Pakwach, 14.3% (156/1094) in Buliisa, and 9.9% (77/776) in Mayuge. Other abnormalities detected were ascites and portosystemic collaterals (gastrooesophageal, pancreaticoduodenal, splenic varices, recanalized paraumbilical veins and splenorenal shunts). Ascites of any severity presented in 0.21% (7/3302) participants, and portosystemic collateral pathways were identified in 1.21% (40/3302) participants. Sonographers detected fatty-like, hepatitis B-like, and cirrhosis-like livers via POCUS in 1.91% (63/3302), 1.57% (52/3302), and 0.45% (15/3302) participants, respectively. 2.8% (51/1817) female participants were pregnant.

**Table 4:** Liver fibrosis pattern prevalence. Number of occurrences and prevalence of each liver fibrosis pattern, as identified at the field scan.

| **Pattern** | **Pattern description** | **No. participants (N=3302)** | **Prevalence** |
|---|---|---|---|
| A | Normal | 2245 | 67.99% |
| B | Unclear | 5 | 0.15% |
| B0 | Feather streaks | 302 | 9.15% |
| B1a | Starry sky | 273 | 8.27% |
| B1b | Flying saucers | 69 | 2.09% |
| B2 | Spider thickening | 89 | 2.60% |
| C1 | Prominent peripheral rings | 568 | 17.2% |
| C2 | Prominent pipe stems | 530 | 16.05% |
| D | Ruff around portal bifurcation | 158 | 4.78% |
| E | Patches | 60 | 1.82% |
| F | Bird’s claw | 12 | 0.36% |

### Participants sampled for second sonographer reading

Participants with any liver fibrosis patterns (B–F) were eligible for second sonographer reading. Figure 1 depicts our exclusion criteria that resulted in a sample size of N=1040 participants for analysis of inter-observer agreement. 47 participants were excluded in total of the 1087 participants who had liver fibrosis patterns, due to having missing fibrosis pattern images (46/47), incorrect labelling (a label that did not correspond to possible fibrosis patterns, 1 image), or having no patterns assigned at second reading (1/47). Having no patterns assigned at second reading was considered missingness since an A pattern (representing a normal liver) was allowed to be assigned.

**Fig. 1:**
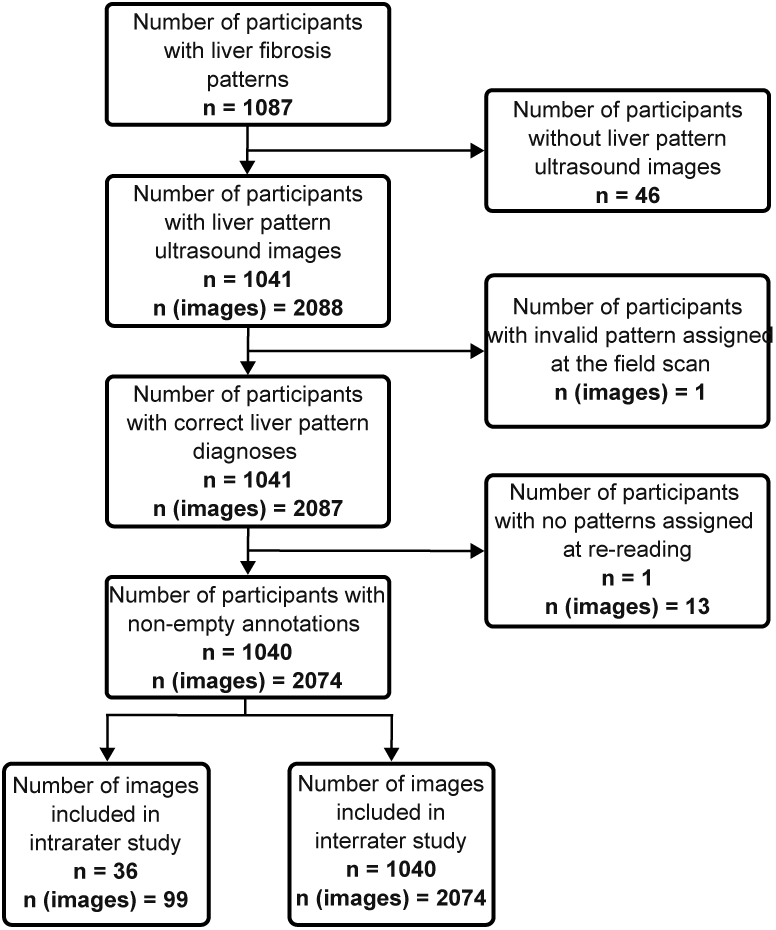
Participant flowchart (images). Flowchart of participants sampled for image analyses. Having an invalid pattern assigned in the field meant there was a label other than B, B0, B1a, B1b, B2, C1, C2, D, E or F outside the field of view.

### Absolute agreement for individual liver fibrosis patterns

Image-level inter-observer agreement analysis was conducted on the set of N=2075 images from 1040 participants. The percentage agreement when considering the granular categories for the B and C patterns was 57.2% (1187/2074). When B (B0, B1a/b and B2) and C (C1 and C2) patterns were combined into single categories, the percentage agreement rose to 73.0% (1515/2074).

The distribution of patterns assigned in the field versus patterns assigned by the second sonographer reading are shown in Figure 2. The distributions follow a similar shape for the C–F patterns for both the field diagnosis and the second sonographer reading. In contrast, the occurrence of A and B patterns was much higher in the second sonographer reading (156 images and 130 images, respectively) compared to the field diagnosis (0 images and 3 images, respectively). Percentage agreement (as assigned in the field) is shown in Tables 5 (a) and 5 (b) for the granular and broader systems respectively. Looking at the granular system, agreement was highest for the C1 and C2 patterns and lowest for the B0, B1a and F patterns. Cohen’s *κ* and ICC for agreement is shown in Table 5 (b). All Cohen’s *κ* and ICC for agreement values were significant (*p <* 0.05). Cohen’s *κ* and ICC for agreement were highest for the D pattern (0.717, *p <* 0.0001 for both *κ* and ICC) and lowest for the F pattern (0.149, *p <* 0.0001 for both *κ* and ICC). ICC for agreement was significantly different between the D pattern and any other pattern, and between the F pattern and any other pattern.

**Fig. 2:**
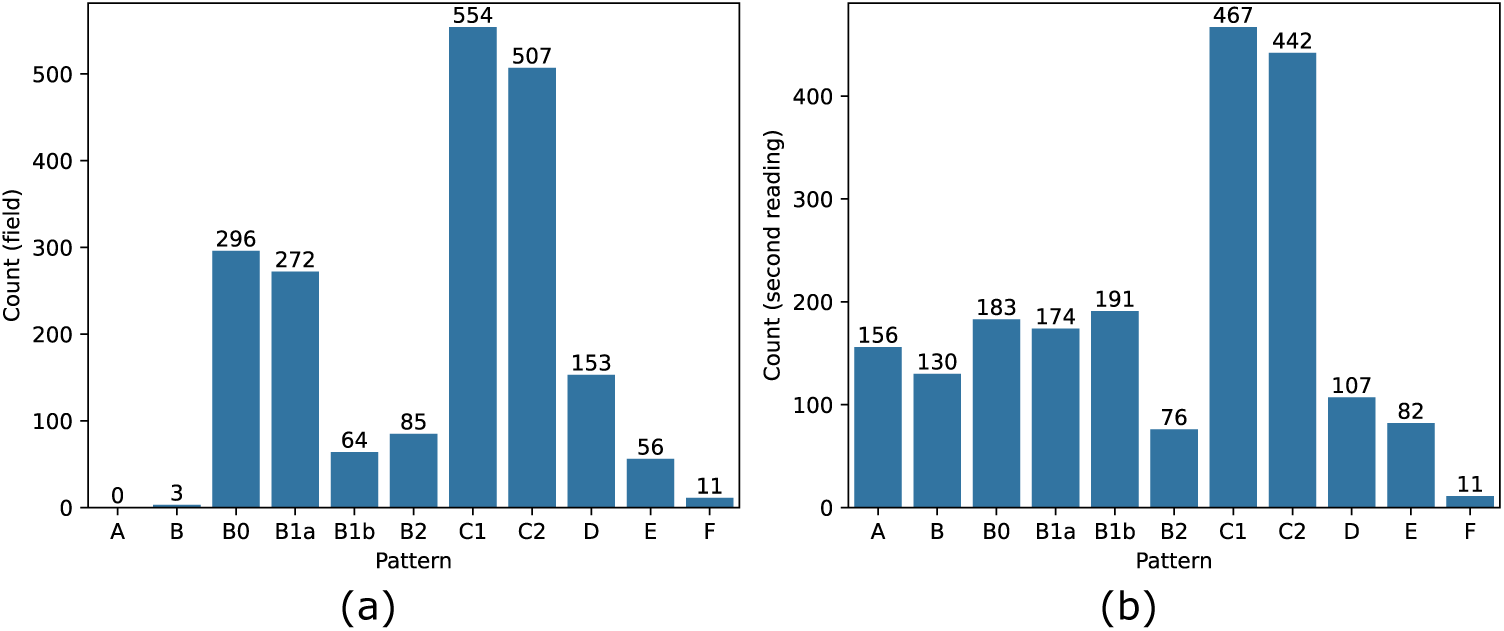
Distributions of liver fibrosis patterns assigned in the field and by the second sonographer. (a) Frequencies of patterns assigned in field scans. (b) Frequencies of patterns assigned in second sonographer reading.

**Table 5:** Percentage agreement with the field pattern as ground truth, using granular patterns. For each pattern, we report the percentage of patterns assigned in the field where the second reading was consistent.

| **Field scan pattern** | **% Agreement** | **Cohen's *κ* (*p*)** | **ICC agreement (*p*) [CI]** |
|---|---|---|---|
| (a) Granular patterns |  |  |  |
| B | 66.67% (2/3) | 0.021 (0.0001) | 0.021 (0.0001) [-0.018, 0.062] |
| B0 | 27.51% (82/298) | 0.224 (< 0.0001) | 0.224 (< 0.0001) [0.183, 0.264] |
| B1a | 34.27% (98/286) | 0.335 (< 0.0001) | 0.335 (< 0.0001) [0.295, 0.373] |
| B1b | 52.17% (36/69) | 0.198 (< 0.0001) | 0.198 (< 0.0001) [0.151, 0.244] |
| B2 | 25.56% (23/90) | 0.238 (< 0.0001) | 0.238 (< 0.0001) [0.197, 0.278] |
| C1 | 73.08% (418/572) | 0.663 (< 0.0001) | 0.663 (< 0.0001) [0.638, 0.686] |
| C2 | 72.95% (383/525) | 0.664 (< 0.0001) | 0.664 (< 0.0001) [0.639, 0.687] |
| D | 65.61% (104/157) | 0.717 (< 0.0001) | 0.717 (< 0.0001) [0.695, 0.738] |
| E | 65.57% (40/61) | 0.429 (< 0.0001) | 0.430 (< 0.0001) [0.393, 0.464] |
| F | 15.38% (2/13) | 0.149 (< 0.0001) | 0.149 (< 0.0001) [0.106, 0.190] |
| (b) Broad patterns |  |  |  |
| B | 72.39% (540/746) | 0.526 (< 0.0001) | 0.526 (< 0.0001) [0.494, 0.556] |
| C | 75.66% (830/1097) | 0.573 (< 0.0001) | 0.573 (< 0.0001) [0.543, 0.602] |
| D | 65.61% (104/157) | 0.717 (< 0.0001) | 0.717 (< 0.0001) [0.695, 0.738] |
| E | 65.57% (40/61) | 0.429 (< 0.0001) | 0.430 (< 0.0001) [0.393, 0.464] |
| F | 15.38% (2/13) | 0.149 (< 0.0001) | 0.149 (< 0.0001) [0.106, 0.190] |

Figure 5 depicts Sankey diagrams by pattern of agreement or disagreement of images from the field to second sonographer reading. For the majority of images, sonographers agreed or only disagreed by one pattern (Figure 3). Correlation between a D pattern during the field scan and a D pattern during the second sonographer reading was strongly positive (0.73, *p <* 0.001) (Figure 4). There were weak positive correlations between the E patterns assigned in the field and F patterns assigned by the second sonographer, and vice versa (0.17, *p <* 0.001 for both). There also was a weak positive correlation (0.17, *p <* 0.001) between the B pattern assigned at in the field and an A pattern assigned by the second sonographer. There were moderate negative correlations between the C patterns from the field and B patterns from the second sonographer (-0.38, *p <* 0.001) and vice versa (-0.26, *p <* 0.001). Supplementary Figures S1 and S2 show a correlation matrix and Sankey diagrams for the granular patterns. Trends were largely similar to the associations found between the broad patterns, in addition significant moderate positive correlations were found between the B0 and B1a patterns being assigned either in the field or at second reading.

**Fig. 3:**
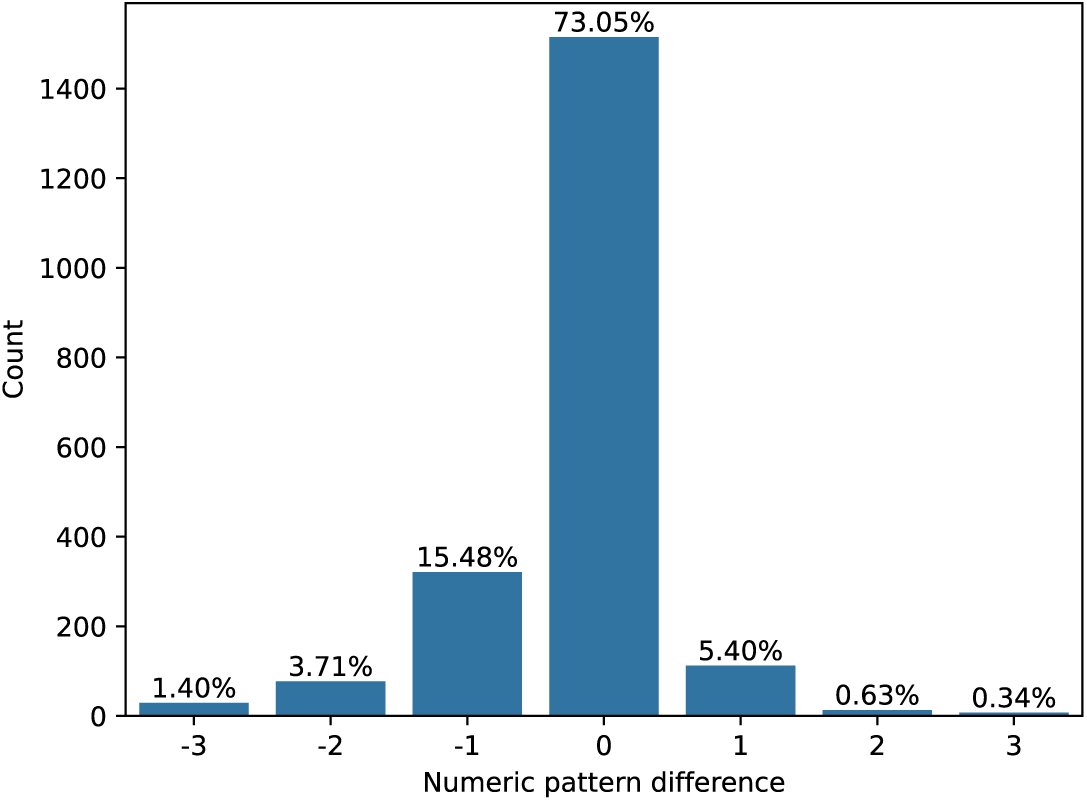
Distribution of numeric pattern differences. To calculate the numeric difference, each pattern was enumerated, where: A=0, B=1, C=2, D=3, E=4, F=5. The difference between the enumerated pattern assigned at the field scan and at the second sonographer reading was the numeric pattern difference.

**Fig. 4:**
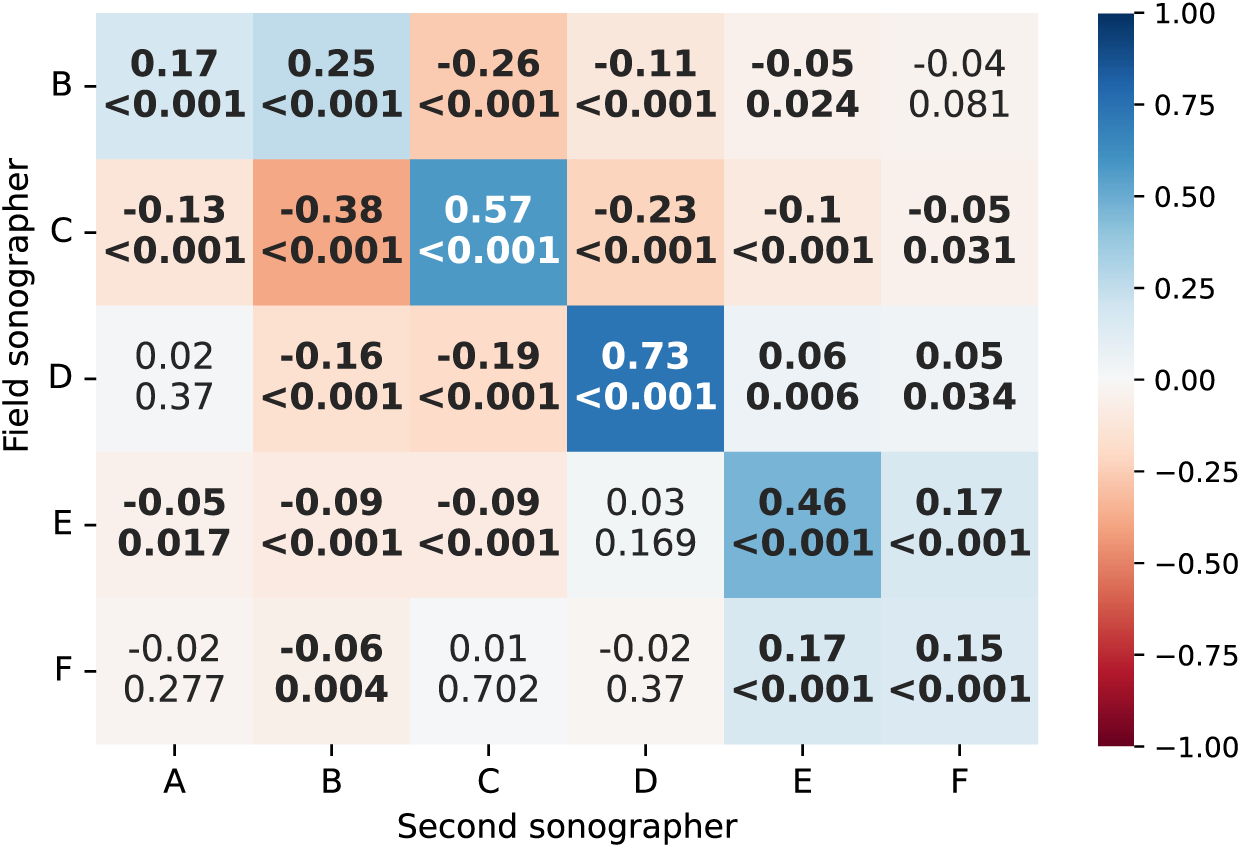
Correlation matrix. Spearman correlation of patterns assigned in the field compared to the second sonographer reading.

**Fig. 5:**
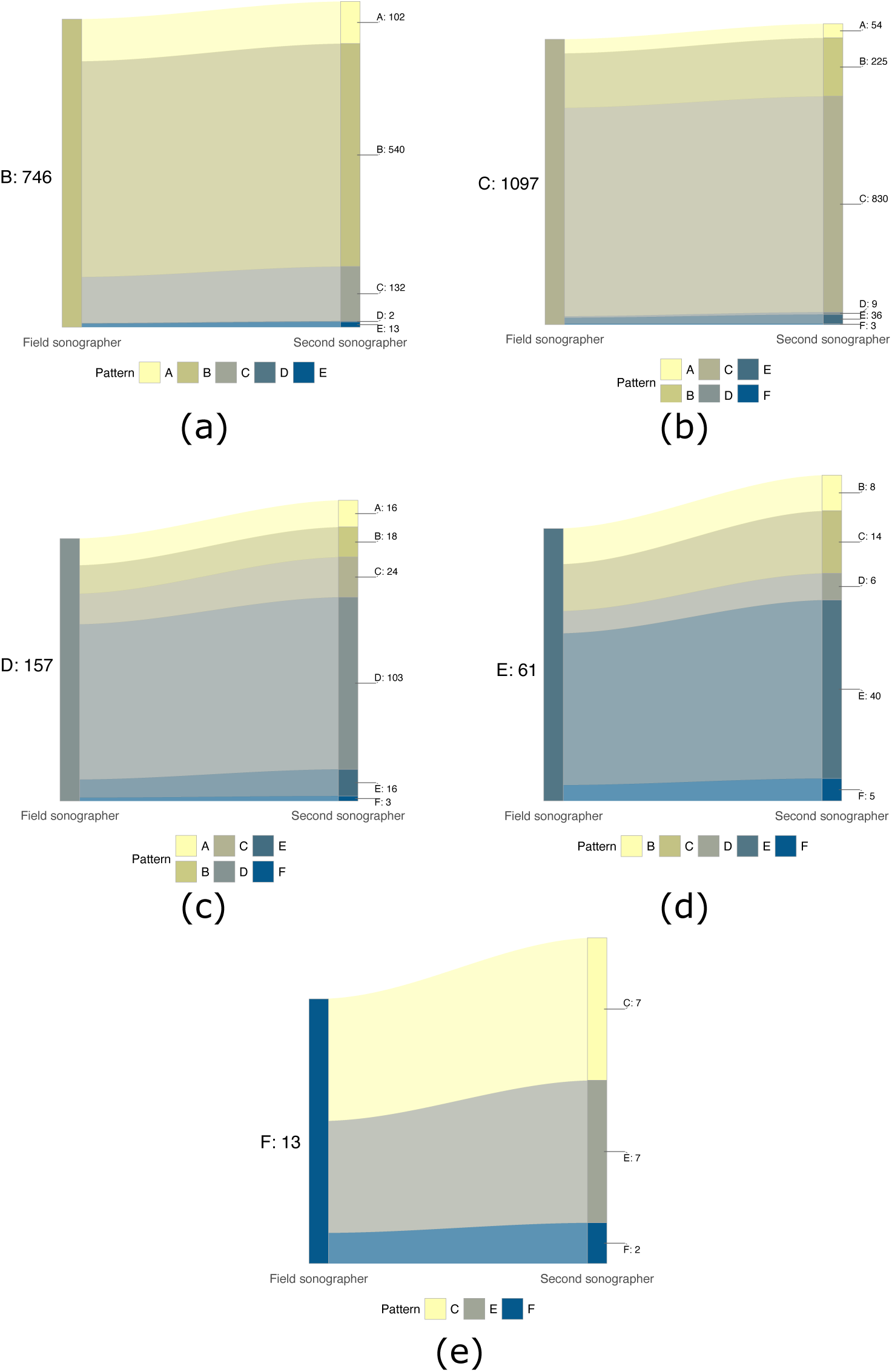
Sankey diagrams of liver pattern diagnoses. Shows patterns assigned at second reading from images of each fibrosis pattern as assigned in the field: (a) B. (b) C. (c) D. (d) E. (e) F.

### Agreement for liver fibrosis severity

Participant-level inter-observer agreement metrics were evaluated using the sample of N=1040 participants. Sonographers agreed on PPF diagnosis for 85.29% (887/1040) of participants overall. Cohen’s *κ* for agreement on PPF diagnosis was 0.701 indicating substantial agreement, and the ICC for agreement was 0.701 [0.669, 0.731] (*p* = 2.87 *×* 10^−155^).

For the highest pattern assigned to a participant (assuming here that the highest pattern was indicative of severity), patterns were grouped into five categories: B, C, D, E and F. The percentage agreement for the highest pattern was 70.29% (731/1040). Cohen’s *κ* was 0.552 indicating moderate agreement, and weighted Cohen’s *κ*, which accounts for class imbalance, was 0.734 indicating substantial agreement. ICC for consistency was 0.128 [0.068,0.167] (*p* = 1.74 *×* 10*^−^*^5^) and ICC for agreement was 0.128 [0.068, 0.187] (*p* = 0.061). For participants who had B as their highest pattern from the field scan, the second sonographer most often assigned an A or C pattern (Figure 6). For participants who had C as their highest pattern during the field scan, the highest patterns from the second sonographer were most often only a B pattern.

**Fig. 6:**
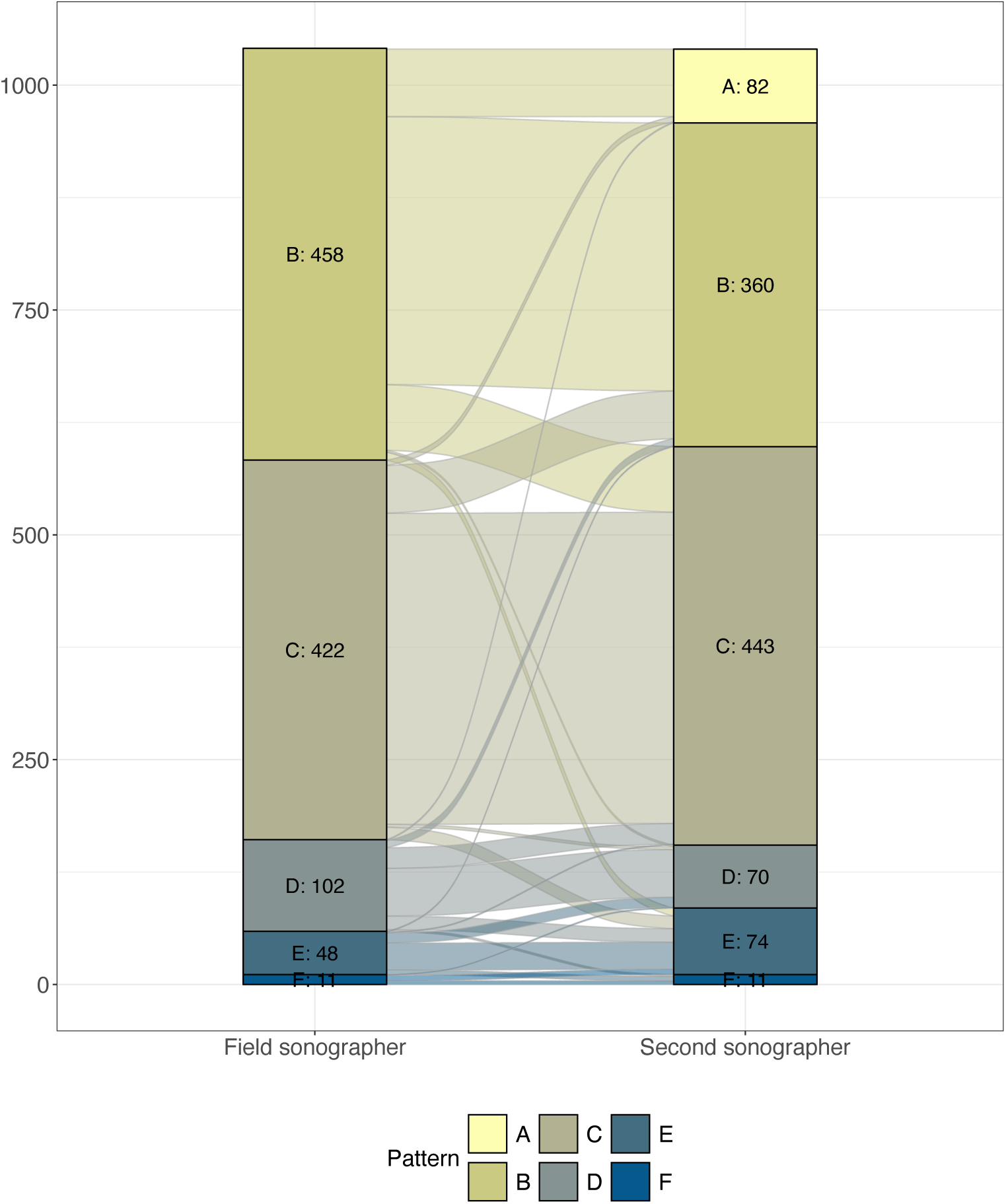
Sankey diagram for highest pattern. Highest pattern was assigned on a participant level, and compared between the field scan to the second sonographer reading.

### Influence of image and participant characteristics on agreement

Image-level and participant-level characteristics were tested against different forms of agreement to assess what characteristics might influence agreement or disagreement. The image-level characteristics were clinical confidence score, probe position, and types of interference which were breathing/movement, gas, bone, fat, calcification and dehydration (Table 6). Calcification was not present in any image. For simple agreement (whether the image was agreed upon by the field and second sonographer according to the B, C, D, E, F grouping), clinical confidence score, probe position, and interference due to breathing/movement and fat were all significant (*p <* 0.05) according to *χ*^2^ tests. Clinical confidence score was significant for all the individual patterns, whereas probe position was only significant (*p <* 0.05) for the C pattern; and breathing/movement and fat were only significant (*p <* 0.05) for the B and C patterns. Table 7 presents a summary of participant characteristics. Situs inversus was not present in any participant. BMI was missing in 8 participants, HIV status was missing in 20 participants, and hepatitis B infection data was missing in 2 participants, leaving 1011 participants for this analysis (missingness was present in more than one column for one participant). Field sonographers did not have the same distribution of patterns assigned in the field; in particular sonographers 1 and 2 scanned most participants with E or F patterns.

**Table 6:**
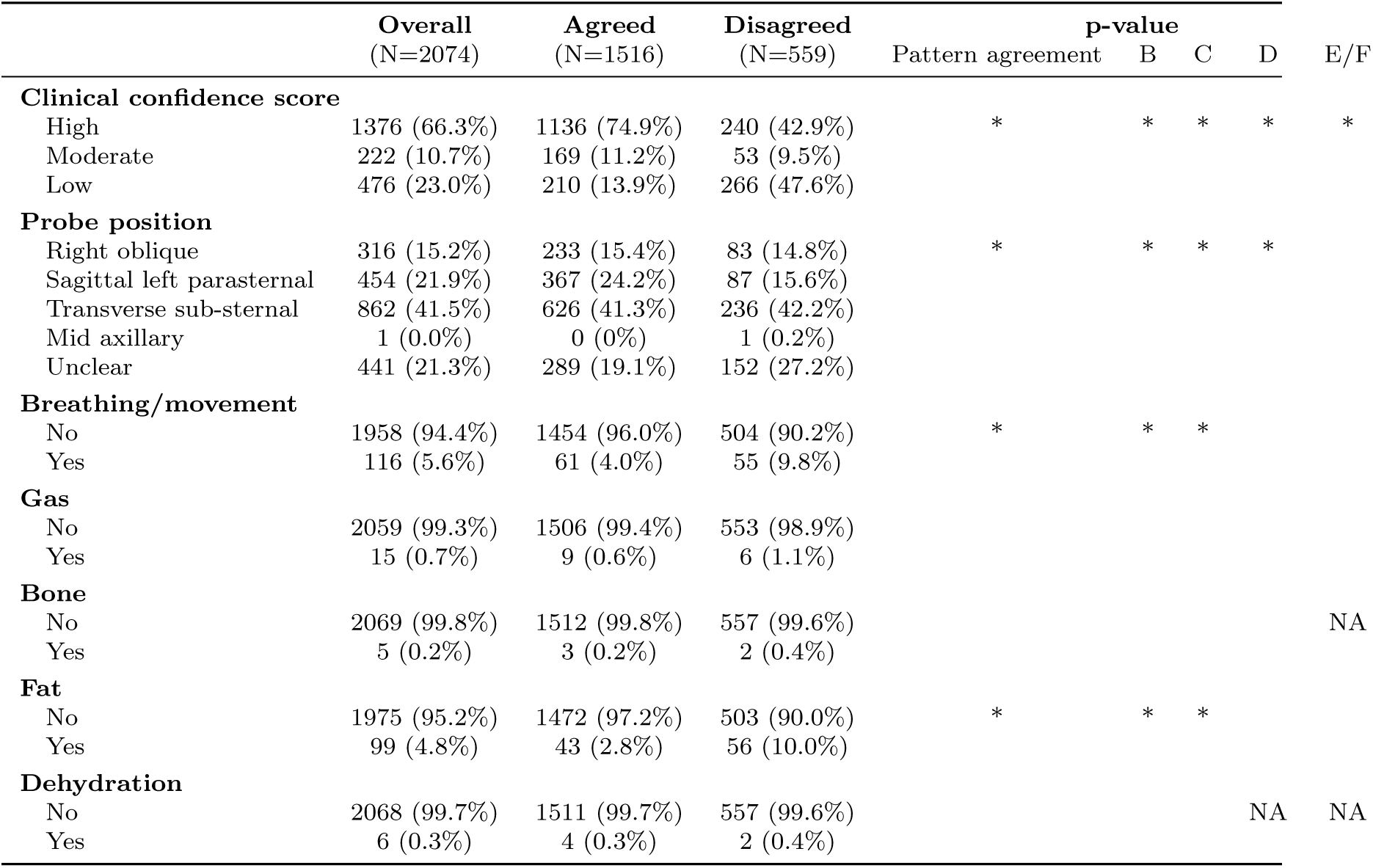
Image-level characteristics. A * in the p-value column indicates a p-value *<* 0.05. NAs are present where there were no examples of that characteristic in images of D or E/F patterns.

|  | **Overall** | **Agreed** | **Disagreed** | **p-value** |  |  |  |  |
|  | (N=2074) | (N=1516) | (N=559) | Pattern agreement | B | C | D | E/F |
|---|---|---|---|---|---|---|---|---|
| **Clinical confidence score** |  |  |  |  |  |  |  |  |
| High | 1376 (66.3%) | 1136 (74.9%) | 240 (42.9%) | * | * | * | * | * |
| Moderate | 222 (10.7%) | 169 (11.2%) | 53 (9.5%) |  |  |  |  |  |
| Low | 476 (23.0%) | 210 (13.9%) | 266 (47.6%) |  |  |  |  |  |
| **Probe position** |  |  |  |  |  |  |  |  |
| Right oblique | 316 (15.2%) | 233 (15.4%) | 83 (14.8%) | * | * | * | * |  |
| Sagittal left parasternal | 454 (21.9%) | 367 (24.2%) | 87 (15.6%) |  |  |  |  |  |
| Transverse sub-sternal | 862 (41.5%) | 626 (41.3%) | 236 (42.2%) |  |  |  |  |  |
| Mid axillary | 1 (0.0%) | 0 (0%) | 1 (0.2%) |  |  |  |  |  |
| Unclear | 441 (21.3%) | 289 (19.1%) | 152 (27.2%) |  |  |  |  |  |
| **Breathing/movement** |  |  |  |  |  |  |  |  |
| No | 1958 (94.4%) | 1454 (96.0%) | 504 (90.2%) | * | * | * |  |  |
| Yes | 116 (5.6%) | 61 (4.0%) | 55 (9.8%) |  |  |  |  |  |
| **Gas** |  |  |  |  |  |  |  |  |
| No | 2059 (99.3%) | 1506 (99.4%) | 553 (98.9%) |  |  |  |  |  |
| Yes | 15 (0.7%) | 9 (0.6%) | 6 (1.1%) |  |  |  |  |  |
| **Bone** |  |  |  |  |  |  |  |  |
| No | 2069 (99.8%) | 1512 (99.8%) | 557 (99.6%) |  |  |  |  | NA |
| Yes | 5 (0.2%) | 3 (0.2%) | 2 (0.4%) |  |  |  |  |  |
| **Fat** |  |  |  |  |  |  |  |  |
| No | 1975 (95.2%) | 1472 (97.2%) | 503 (90.0%) | * | * | * |  |  |
| Yes | 99 (4.8%) | 43 (2.8%) | 56 (10.0%) |  |  |  |  |  |
| **Dehydration** |  |  |  |  |  |  |  |  |
| No | 2068 (99.7%) | 1511 (99.7%) | 557 (99.6%) |  |  |  | NA | NA |
| Yes | 6 (0.3%) | 4 (0.3%) | 2 (0.4%) |  |  |  |  |  |

**Table 7:**
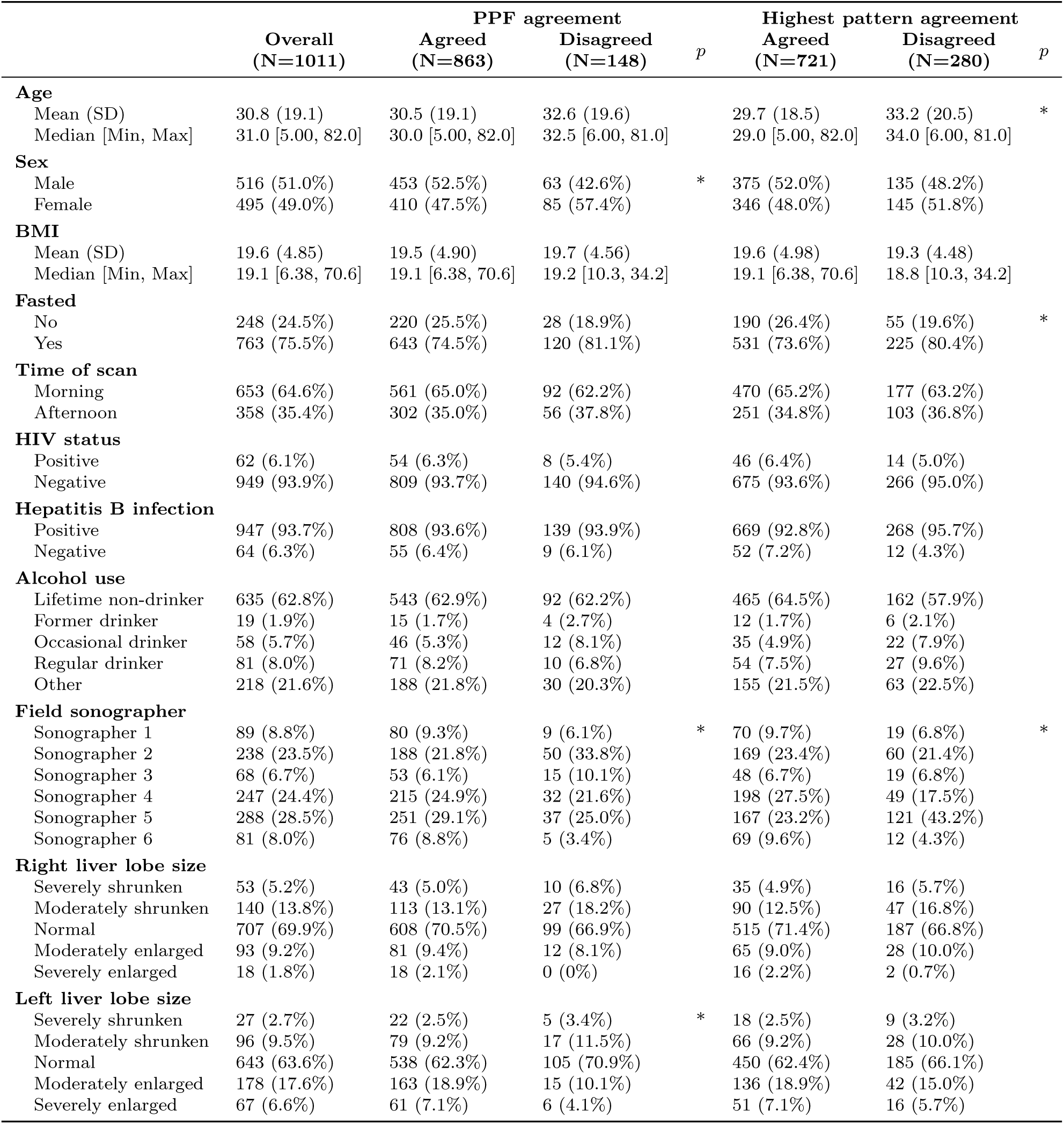
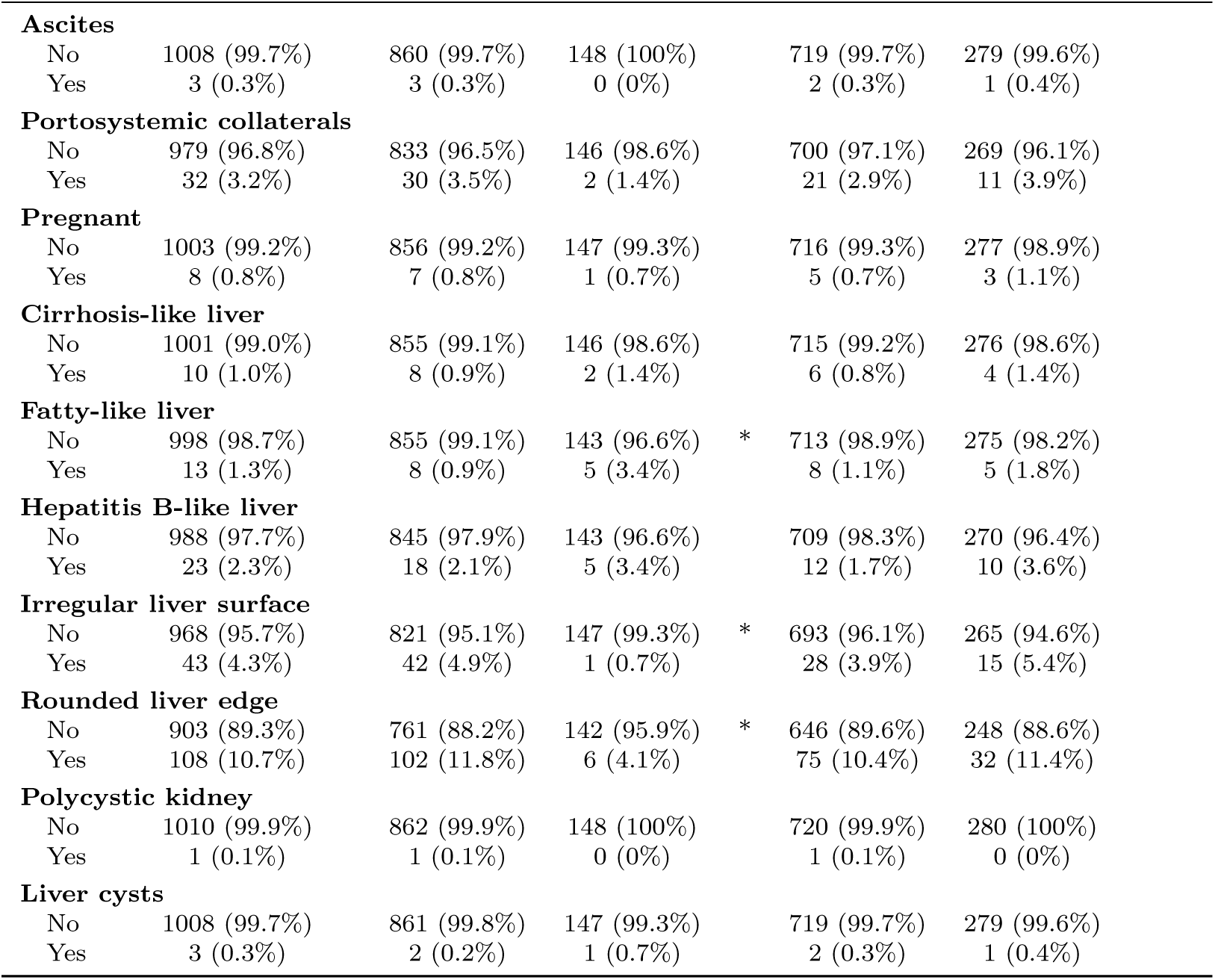
Participant-level characteristics. A * in the p-value column indicates a p-value *<* 0.05.

Age was significant (*p* = 0.012) when looking at highest pattern agreement, with older participants being more likely to have disagreement, but not for PPF agreement (*p* = 0.223); with female sex being significant for PPF agreement (*p* = 0.032) but not highest pattern agreement (*p* = 0.313). Fasting status was significant (*p* = 0.033) for the highest pattern agreement, where fasted participants were more likely to have disagreement but not for PPF agreement (*p* = 0.107). The sonographer that performed the field scan was significant for both PPF agreement (*p* = 0.003) and highest pattern agreement (*p <* 0.001). Left liver lobe size was significant for PPF agreement (*p* = 0.045) but not for highest pattern agreement (*p* = 0.523). None of the sonographical findings were significant for highest pattern agreement, but fatty liver (*p* = 0.040), irregular liver surface (*p* = 0.035) and rounded liver edge were significant (*p* = 0.007) for PPF agreement. Participants with fatty-like liver were more likely to have disagreement, where those with an irregular liver surface and rounded liver edge were less likely to have disagreement.

### Agreement using ultrasound video

The number of participants whose videos were sent for second reading was 1060. The number of participants whose sweep one video was clear enough to read for fibrosis patterns was 1054 (Figure 7). The percentage agreement between the field sonographer and the second sonographer for the exact pattern of fibrosis was 31.2% (329/1054) and when granular B patterns and C patterns were grouped the percentage agreement was 43.6% (460/1054). The percentage agreement for each granular pattern is shown in Table 8 (a) and with grouped Bs and Cs in Table 8 (b). Cohen’s *κ* was *<* 0.1 for all patterns, indicating no agreement and the ICC for agreement was also *<* 0.1 for every pattern (Table 8 (b)). Only the E pattern had *p <* 0.05 for Cohen’s *κ* and ICC for agreement.

**Fig. 7:**
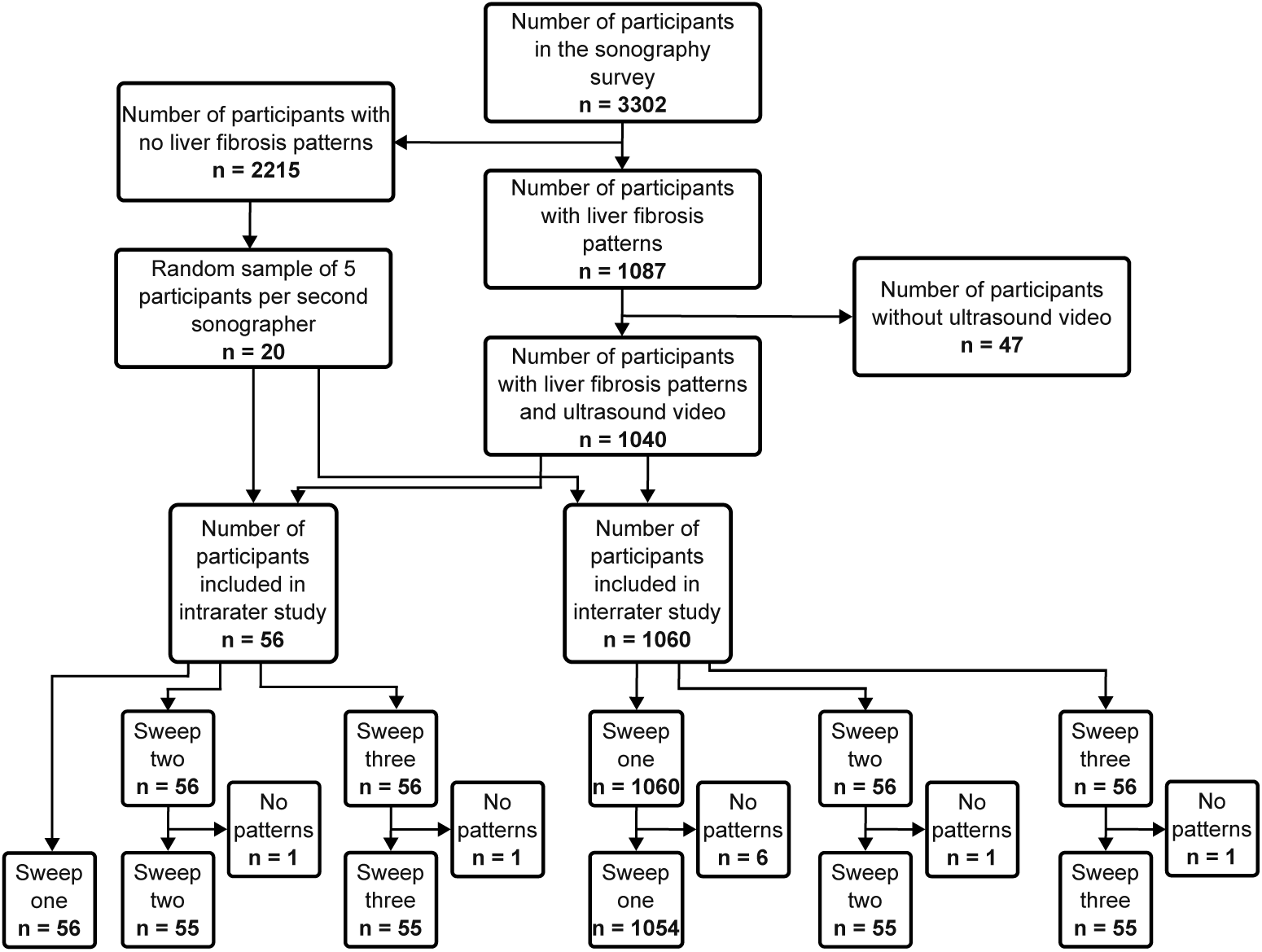
Participant flowchart (videos). Flowchart of participants used for ultrasound video analysis.

**Table 8:** Agreement for ultrasound video. For Cohen’s *κ* and ICC for agreement, *p*-values are given in parentheses. 95% confidence intervals (CIs) for ICC for agreement are given in square brackets. Statistics are given for (a) granular patterns and (b) broad patterns.

| **Field scan pattern** | **% Agreement** | **Cohen's *κ* (*p*)** | **ICC agreement (*p*) [CI]** |
|---|---|---|---|
| (a) Granular patterns |  |  |  |
| A | 40.00% (8/20) | 0.016 (0.228) | 0.016 (0.257) [-0.031, 0.063] |
| B | 33.33% (1/3) | 0.037 (0.011) | 0.037 (0.104) [-0.021, 0.096] |
| B0 | 38.10% (112/294) | 0.093 (0.002) | 0.093 (0.001) [0.033, 0.152] |
| B1a | 16.42% (44/268) | 0.028 (0.328) | 0.028 (0.170) [-0.030, 0.086] |
| B1b | 7.94% (5/63) | -0.009 (0.759) | -0.009 (0.618) [-0.069, 0.051] |
| B2 | 2.35% (2/85) | 0.011 (0.626) | 0.011 (0.353) [-0.046, 0.069] |
| C1 | 19.82% (109/550) | -0.035 (0.148) | -0.035 (0.907) [-0.087, 0.017] |
| C2 | 28.03% (141/503) | -0.033 (0.240) | -0.033 (0.877) [-0.090, 0.023] |
| D | 4.58% (7/153) | 0.022 (0.341) | 0.022 (0.221) [-0.034, 0.078] |
| E | 7.14% (4/56) | 0.062 (0.031) | 0.062 (0.020) [0.003, 0.122] |
| F | 0.0% (0/11) | -0.007 (0.818) | -0.007 (0.584) [-0.067, 0.054] |
| (b) Broad patterns |  |  |  |
| A | 40.00% (8/20) | 0.016 (0.228) | 0.016 (0.257) [-0.031, 0.063] |
| B | 49.15% (289/588) | 0.041 (0.140) | 0.045 (0.070) [-0.015, 0.104] |
| C | 31.31% (175/559) | -0.025 (0.381) | -0.025 (0.807) [-0.080, 0.031] |
| D | 4.58% (7/153) | 0.022 (0.341) | 0.022 (0.221) [-0.034, 0.078] |
| E | 7.14% (4/56) | 0.062 (0.031) | 0.062 (0.020) [0.003, 0.122] |
| F | 0.0% (0/11) | -0.007 (0.818) | -0.007 (0.584) [-0.067, 0.054] |

For video reading, sonographers agreed on PPF diagnosis for 48.86% (515/1054) of participants using the sweep one video. Cohen’s *κ* for agreement on PPF diagnosis was 0.001 indicating no agreement, and the ICC for agreement was 0.001 [-0.055, 0.058] (*p* = 0.480). For the agreement on the highest pattern using the same groupings as the participant level analysis, Cohen’s *κ* was 0.009, and weighted Cohen’s *κ*, which accounts for class imbalance, was 0.037. ICC for consistency was -0.008 [-0.068, 0.053] (*p* = 0.598) and ICC for agreement was 0.003 [-0.056, 0.062] (*p* = 0.465).

To assess whether the lack of agreement in videos was due to the scanning style, we conducted an intra-observer study to assess whether sonographers can better reread their own videos. 56 participants were included with sweep one videos for the intraobserver study (Figure 7). The percentage agreement for any pattern was 37.5% (21/56) when the Bs and Cs were grouped, which was lower than for the inter-observer study of videos, and the percentage PPF agreement was 57.1% (32/56) which was higher than for the inter-observer study of videos. The ICC for agreement was 0.087 [-0.183, 0.343] (*p* = 0.263) for PPF and was 0.005 [-0.251, 0.262] (*p* = 0.485) for agreement on the highest pattern. To assess whether variability was mainly due to videos, we also conducted a intra-observer study for images. For the intra-observer study of images, agreement was higher than that observed for video. 63.6% (63/99) images were agreed upon between the field and second sonographers for the exact pattern, and when the Bs and Cs were grouped the percentage agreement was 81.8% (81/99). Sweep two and three videos from 56 participants to be read by a second sonographer (Table 7). The sweep used to assess agreement for each pattern was important. We analysed agreement on a subset of videos for sweep two and three. For sweep two, which targets the main portal vein, the percentage agreement for the exact pattern when the Bs and Cs were grouped was 65.5% (31/55). For the D pattern, which is found around the main portal vein, the percentage agreement was 50.0% (5/10) which was much higher than the agreement for the D pattern in sweep one video (4.58%, 7/153). PPF diagnosis was agreed upon for 78.1% (43/55) of participants. Cohen’s *κ* and ICC for agreement for PPF diagnosis were both higher for sweep two than for sweep one (*κ*: 0.001 compared to 0.490 and ICC: 0.001 compared to 0.495) (see Table 9 (a)). For highest pattern agreement, Cohen’s *κ* and Cohen’s *κ_W_* were higher for sweep two than sweep one, but there were no differences in ICCs for agreement and consistency between sweep one and two (see Table 9 (b)).

**Table 9:**
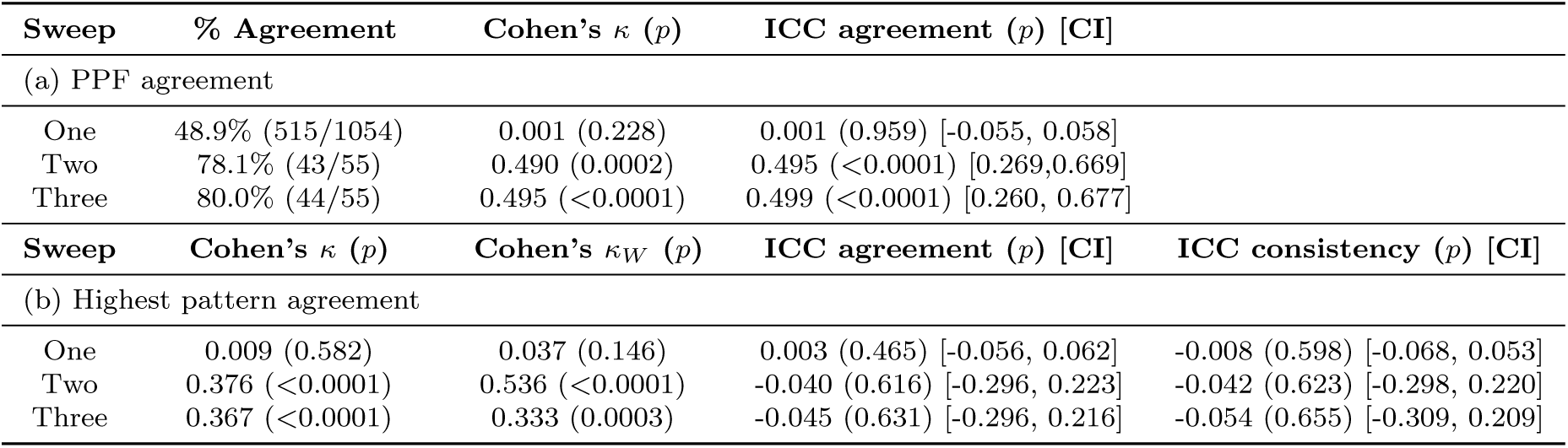
Percentage agreement, *κ* and ICCs for video sweeps. We report the (a) percentage agreement, Cohen’s *κ* and ICC for agreement (with 95% CIs) for PPF agreement and (b) Cohen’s *κ*, *κ_W_*, ICC for agreement (with 95% CIs) and ICC for consistency (with 95% CIs) for highest pattern agreement. All statistics are for the inter-rater study.

| **Sweep** | **% Agreement** | **Cohen's κ (*p*)** | **ICC agreement (*p*) [CI]** |  |
|---|---|---|---|---|
| (a) PPF agreement |  |  |  |  |
| One | 48.9% (515/1054) | 0.001 (0.228) | 0.001 (0.959) [-0.055, 0.058] |  |
| Two | 78.1% (43/55) | 0.490 (0.0002) | 0.495 (<0.0001) [0.269,0.669] |  |
| Three | 80.0% (44/55) | 0.495 (<0.0001) | 0.499 (<0.0001) [0.260, 0.677] |  |
| **Sweep** | **Cohen's κ (*p*)** | **Cohen's κ<sub>*W*</sub> (*p*)** | **ICC agreement (*p*) [CI]** | **ICC consistency (*p*) [CI]** |
| (b) Highest pattern agreement |  |  |  |  |
| One | 0.009 (0.582) | 0.037 (0.146) | 0.003 (0.465) [-0.056, 0.062] | -0.008 (0.598) [-0.068, 0.053] |
| Two | 0.376 (<0.0001) | 0.536 (<0.0001) | -0.040 (0.616) [-0.296, 0.223] | -0.042 (0.623) [-0.298, 0.220] |
| Three | 0.367 (<0.0001) | 0.333 (0.0003) | -0.045 (0.631) [-0.296, 0.216] | -0.054 (0.655) [-0.309, 0.209] |

For sweep three, which targets sagittal views of the left liver lobe, the percentage agreement for the exact pattern when the Bs and Cs were grouped was 72.7% (40/55). For the C1 pattern, the percentage agreement was 52.9% (9/17) compared to 19.8% (109/550) for sweep one. PPF diagnosis was agreed upon for 80% (44/55) of participants. Cohen’s *κ* and ICC for agreement for PPF diagnosis were both higher for sweep three than for sweep one (*κ*: 0.001 compared to 0.495 and ICC: 0.001 compared to 0.499) (see Table 9 (a)). For highest pattern agreement, Cohen’s *κ* and Cohen’s *κ_W_* were higher for sweep three than sweep one, but there were no differences in ICCs for agreement and consistency between sweep one and two (see Table 9 (b)).

The characteristics for the videos included for the inter-observer analysis are shown in Table 10. The confidence and interference types were the same as for the image analysis, and the sonographers recorded whether the sweep procedure was followed. There were no videos that had calcification present. For sweep two and three, the percentage of videos with high confidence was lower than for sweep one (23.6% and 25.5% compared to 52.6%). The majority of videos followed the sweep procedure for all three sweeps (92.8%, 978/1054 for sweep one; 87.3%, 42/55 for sweep two; and 94.5%, 52/55 for sweep three). Interference caused by breathing/movement and by gas was present mostly in sweep two (gas present in 80.0%, 44/55 videos; breathing/movement in 76.4%, 42/55 videos) and three videos (gas present in 78.2%, 43/55 videos; breathing/movement in 76.4%, 42/55 videos) but in few sweep one videos (gas present in 21.4%, 226/1054 videos; breathing/movement in 12.3%, 130/1054 videos).

**Table 10:** Video characteristics. A summary of the prevalence of clinical confidence scores, whether the sweep procedure was followed, and types of interference among the ultrasound videos that were read by a second sonographer.

|  | **Sweep one (N=1054)** | **Sweep two (N=55)** | **Sweep three (N=55)** |
|---|---|---|---|
| **Clinical confidence score** |  |  |  |
| High | 554 (52.6%) | 13 (23.6%) | 14 (25.5%) |
| Moderate | 368 (34.9%) | 30 (54.5%) | 34 (61.8%) |
| Low | 132 (12.5%) | 12 (21.8%) | 7 (12.7%) |
| **Sweep procedure followed** |  |  |  |
| No | 76 (7.2%) | 7 (12.7%) | 3 (5.5%) |
| Yes | 978 (92.8%) | 48 (87.3%) | 52 (94.5%) |
| **Breathing/movement** |  |  |  |
| No | 924 (87.7%) | 13 (23.6%) | 13 (23.6%) |
| Yes | 130 (12.3%) | 42 (76.4%) | 42 (76.4%) |
| **Gas** |  |  |  |
| No | 828 (78.6%) | 11 (20.0%) | 12 (21.8%) |
| Yes | 226 (21.4%) | 44 (80.0%) | 43 (78.2%) |
| **Bone** |  |  |  |
| No | 1046 (99.2%) | 37 (67.3%) | 35 (63.6%) |
| Yes | 8 (0.8%) | 18 (32.7%) | 20 (36.4%) |
| **Fat** |  |  |  |
| No | 1006 (95.4%) | 49 (89.1%) | 51 (92.7%) |
| Yes | 48 (4.6%) | 6 (10.9%) | 4 (7.3%) |
| **Dehydration** |  |  |  |
| No | 1043 (99.0%) | 52 (94.5%) | 51 (92.7%) |
| Yes | 11 (1.0%) | 3 (5.5%) | 4 (7.3%) |

## Discussion

There is an urgent need to update the WHO Niamey protocol to enable morbidity surveillance and integration of diagnostics for schistosomal morbidity within primary healthcare facilities. Here we conducted a large-scale study on inter-observer agreement for schistosomal liver fibrosis outcomes assessed using ultrasound imaging. We accounted for detailed information related to liver fibrosis pattern and their granularity, assessed both static images and video, surveyed participant characteristics, recorded image quality and sonographer clinical confidence as well as accounted for any other ultrasound abnormalities. We studied 3302 participants from rural districts of Uganda to assess for periportal fibrosis (PPF) diagnosis, and sent all positive cases for any type of fibrosis for reading by a second sonographer. We found that agreement was lowest for E and F patterns and highest for the D pattern from static images with little to no agreement observable for ultrasound video.

Inter-observer agreement is an important aspect of protocol validation, and was presented here to show how our modifications to the WHO Niamey protocol improved standardisation [46, 47]. The results were encouraging, with inter-observer agreement on PPF diagnosis found to be 85.29% overall, and Cohen’s *κ* of 0.701 constituting substantial agreement. Specific acquisition guidance for each fibrosis pattern was agreed upon among Ugandan sonographers and members of the SchistoTrack study team in Oxford. The study findings suggest that our protocol is repeatable and offers a reliable method of diagnosing PPF using POCUS.

Our findings suggest that a continuous scale for staging fibrosis may be more appropriate for schistosomal PPF than discrete patterns. The ultrasound image and video collection protocol used for this study was based on the WHO Niamey protocol [6], which proposed six discrete schistosomal liver fibrosis patterns. For our study, 94.0% of second sonographer readings were within one step of the field sonographer diagnosis. This could be due to ambiguities between patterns of similar type or severity, where it is unclear when exactly a pattern becomes the next. However, introducing more patterns causes more disagreement according to our granular pattern analysis, therefore a continuous system for fibrosis presentations may be preferable and requires future research. To establish and validate a continuous fibrosis scale for incorporation into revisions of the WHO Niamey protocol, studies should be done to link a continual measure of liver fibrosis as well as the existing discrete liver fibrosis patterns to long term prospective clinical outcomes to better understand their relative severity. Alternative forms of image analysis should be considered beyond visual inspection such as continuous fibrosis diagnosis aided through machine learning, possibly aided by transient elastography.

The WHO Niamey protocol recommends that patients fast for four hours prior to ultrasound examination, and that schistosomiasis-related liver patterns should not be diagnosed in cases of fatty liver disease [6]. Our results were not conclusive on the effect of fasting or fatty liver disease on disagreement, since fasting was only significant for highest pattern agreement and fatty liver was only significant for PPF agreement. However, there were few cases of fatty liver and so further studies may be needed to confirm the effect of this. Fasting before abdominal ultrasound examination is standard guidance, since it reduces gas in the abdomen and enables clearer visualisation of organs. However, there is a growing body of literature, including two randomised controlled trials, that show that there is little effect of fasting on ultrasound image quality [48–51]. These studies took place in medical facilities and used cart-based ultrasound machines, so more evidence is needed to determine the effect of fasting on ultrasound image quality in rural, community-based settings. Fatty liver disease may also occur in populations with high schistosomiasis burden due to alcohol consumption, though the interaction between the two is not fully understood [52]. Therefore, recommending that schistosomal liver fibrosis patterns should be ignored in cases of fatty liver could lead to underestimates of disease burden. Our study shows that sonographers agree on the highest pattern of liver fibrosis even when fatty liver disease is present, so this recommendation should be reconsidered in future versions of the WHO Niamey protocol.

Our study compares diagnoses that sonographers made while scanning a participant and diagnoses based on a single static image. Reading of a single image by a second sonographer not involved in scanning has been used in clinical trials as the outcome [53] and therefore has been suggested as a reference standard. However, our study shows that there is variation between the diagnosis given in the field compared to the one given at second reading. The field reading is done in the presence of the participant after a scoping scan, and so the sonographer has much more information on which to base their diagnosis which may make it more reliable. However, this information may also induce bias, for example if there are no clinical signs such as a distended abdomen despite severe fibrosis being present. This imbalance in information means that some of the disagreement could be attributed to disagreement between sonographers, and some due to lack of information. Our intra-observer study shows little difference in agreement when sonographers re-read their own images and there were not many participant characteristics that made a difference to agreement, meaning that the imbalance in information is likely to be the reason why disagreement persists. The field sonographer did have a significant effect on disagreement; however it cannot be discerned whether this is due to scanning style, experience, or that sonographer being assigned harder cases in the field. Future work should conduct larger-scale intra-rater studies and look at intra-rater agreement variation between sonographers with different amounts of experience. A study done in Ethiopia shows very high Cohen’s *κ* values (0.89) when sonographers re-scanned participants rather than re-read images [17]. Therefore, we suggest that the reference standard for PPF diagnosis should be the diagnosis given at the point of image acquisition.

Though our study shows strong agreement for any PPF, there were varying levels of agreement for the specific C–F patterns. In particular, the agreement for the severe (E and F) patterns was lower than for the C and D patterns. The disagreement tended to be one step from the field diagnosis, and were more likely to be an underdiagnosis than an overdiagnosis. Though this minimal difference is encouraging, one step in severity difference may have vastly different implications for clinical management [4]. For example, it is assumed that a one step difference can change the risk of oesophageal varices [6]. Underdiagnosis also has implications for treatment. There is some suggestive evidence that praziquantel can induce upper gastrointestinal bleeding [54], so administration may need to be reconsidered for severe liver fibrosis patterns (E/F). However, praziquantel treatment is recommended in mild and moderate cases of PPF to clear any current infection, and there is some incomplete evidence to suggest that it might reverse mild fibrosis patterns [55]. For outcomes evaluated in epidemiological studies and clinical trials, underdiagnosis could mean overestimation of treatment effects and underestimation of morbidity burden. To counter this, in clinical practice, it may be appropriate to re-scan patients that present with severe patterns or to provide additional clinical signs to give a more thorough diagnosis. In settings endemic with schistosomiasis, it may be challenging to get a second scanner for confirmatory diagnoses, and so establishing and validating a reference standard (the field scanner) would provide clarity on diagnoses for severe cases.

The second sonographer agreed far less with the field sonographer when video sweeps were read rather than ultrasound images. When only sweep one was considered, there was no PPF agreement according to Cohen’s *κ*, with less than half of participants being agreed upon with respect to PPF status. Cohen’s *κ* was very low for the individual patterns, sometimes representing second reading agreeing with the field reading less than chance for sweep one videos. However, for sweep two and three videos the percentage agreement was higher for the D and C1 patterns respectively. Although the sweep one videos showed little agreement, the sonographers were able to pick up the targeted anatomies in the second and third sweeps. When training for diagnosis of periportal fibrosis, sonographers acquire and read images and not videos, so future training should focus on how to interpret video that is not from a live scan. There may also be issues with playback and the quantity of information in a video. These issues might partially explain the lower agreement for ultrasound video. For other applications, for example for breast ultrasound, it has been found that the diagnostic sensitivity and specificity was the same or better when sonographers were reading video rather than images, contrary to our findings [56, 57]. Ultrasound video offers benefits over ultrasound imaging, for example it has been found to be less operator-dependent and easier to standardise [58]. In LMIC settings, disease-specific POCUS protocols proposing simplified ultrasound video sweeps are relatively common, though they have rarely been implemented in clinical practice [59]. The training needed to capture video sweeps, given that they are nonspecific to schistosomiasis morbidity, is far less than that to acquire the liver patterns. Also, the images are of curated views of fibrosis patterns, which may introduce bias because certain patterns are found in certain views, so if a sonographer recognises a view they may assign a particular pattern even if they do not see fibrosis. However, since videos do not follow pathology there is no view-related bias, potentially making videos a better comparator for second sonographer reading though it is a harder task. Future studies should consider the utility of standardised video sweeps for schistosomal PPF and consider training for non-expert sonographers. Videos have been shown to have some utility for PPF diagnosis through the recent development of deep learning-based models [60]. Additional research is needed before recommending the inclusion of videos from the WHO Niamey protocol updates.

There are some limitations to our study. Our second sonographer focused exclusively on participants with fibrosis (B–F), as identified at the field scan, thus we lack inter-observer agreement data for individuals diagnosed with no liver fibrosis patterns in the field. However, we did perform second reading on videos of healthy livers and found no agreement in line with all video-based results. In addition, all participants in our study were from one country and were scanned using the same ultrasound machine. It is possible that populations from different geographies, such as in South America, may present varying challenges in *S. mansoni* -related PPF, and potentially yield different results in inter-sonographer agreement.

## Conclusions

There is an urgent need to monitor schistosomiasis-associated morbidity and to provide clear diagnoses for follow-up and case management in endemic settings. Here we tested our standardised ultrasound protocols that showed good agreement on PPF diagnosis; found that increasing granularity in the assigned patterns had a negative effect on agreement; and found no clear effect of fasting or other image and participant-level factors on agreement. The implications of our findings for clinical care include that agreement for E and F (the most severe patterns) was low, indicating that severe cases may need routine second reading and discussion among experts. For the Niamey protocol, we propose that a continuous scale of fibrosis should be considered and validated, and that more studies should quantify the effect of fatty liver disease on PPF diagnosis.

## Data availability

Image data are protected and are not available due to data privacy laws, restrictions in ethics approvals and participant agreements associated with the ongoing SchistoTrack Cohort study. These restrictions prevent open sharing of raw images and individuallevel participant information that may identify individuals.

## Code availability

Code will be made available in a public repository.

## List of Abbreviations

BMI: body mass index
CI: confidence interval
DICOM: Digital Imaging and COmmunications in Medicine
HIV: human immunodeficiency virus
ICC: intra-class correlation coefficient
JPEG: Joint Photographic Experts Group
LMIC: low- and middle-income country
MP4: Moving Picture Experts Group-4
NTD: neglected tropical disease
ODK: Open Data Kit
POCUS: point-of-care ultrasound imaging
PPF: periportal fibrosis
s.d.: standard deviation
SD: Secure Digital
SOP: standard operating procedure
USB: universal serial bus
WHO: World Health Organization

## Acknowledgments

We are thankful for the involvement of our study participants and the SchistoTrack teams, especially the sonographers, nurses and surveyors. We would also like to thank the Uganda Ministry of Health, local district leaders, focal health workers, and village health teams. We are grateful for the constant involvement through the study time-points and community engagement meetings of our study participants. Special thanks also to the SchistoTrack Group in Oxford for everyday discussions and feedback.

## Author contributions

Conceptualisation: GFC. Data curation: EO, CH, VA, SM, TM, BNt, RN, IT, BNa and GFC. Formal analysis: CH and EO. Funding acquisition: EO and GFC. Investigation and Methodology: CH and GFC. Project administration: BNa, NBK, and GFC. Resources: GFC. Software: GFC. Supervision: GFC. Validation: EO, CH, VA, SM, TM, BNt, RN, IT, and GFC. Visualisation: EO and CH. Writing - original draft: EO and CH. Writing - review & editing: EO, CH, VA, SM, TM, BNt, BNa, RN, IT, NBK and GFC.

## Ethics approval and consent to participate

Data collection and use were reviewed and approved by Oxford Tropical Research Ethics Committee (OxTREC 509-21), Vector Control Division Research Ethics Committee of the Uganda Ministry of Health (VCDREC146), and Uganda National Council of Science and Technology (UNCST HS 1664ES). Written informed consent was obtained from adult participants aged 18 years and older, who also provided written consent on behalf of verbally assented children with fingerprint assent. Older children also provided written consent in addition to the adult consent on their behalf.

