## Supplementary Figures for "Evaluation of inter-observer agreement for periportal fibrosis related to *Schistosoma mansoni*: A population-based study of 3302 individuals using point-of-care ultrasound images and video in the SchistoTrack cohort"

### Appendix S

#### Results using granular patterns

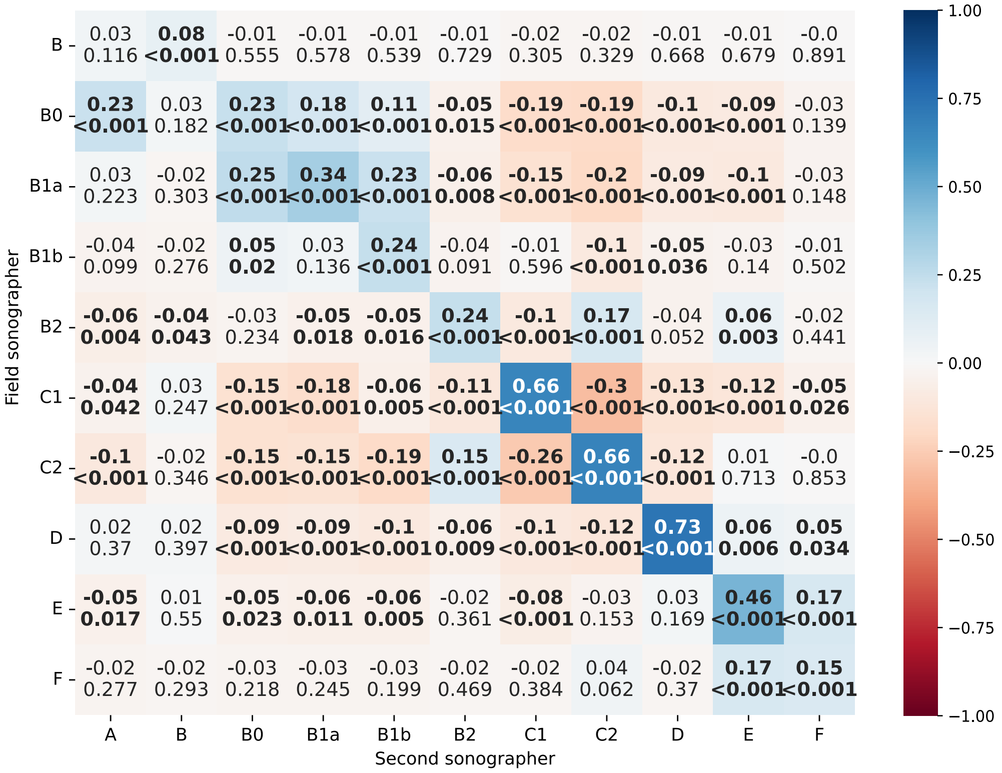


**Fig. S1**: **Correlation matrix.** Correlation of patterns assigned at point-of-care ultrasound imaging (POCUS) vs second sonographer reading, using granular patterns. The top row for the A pattern is blank because there were no images for A patterns assigned at the field scan.

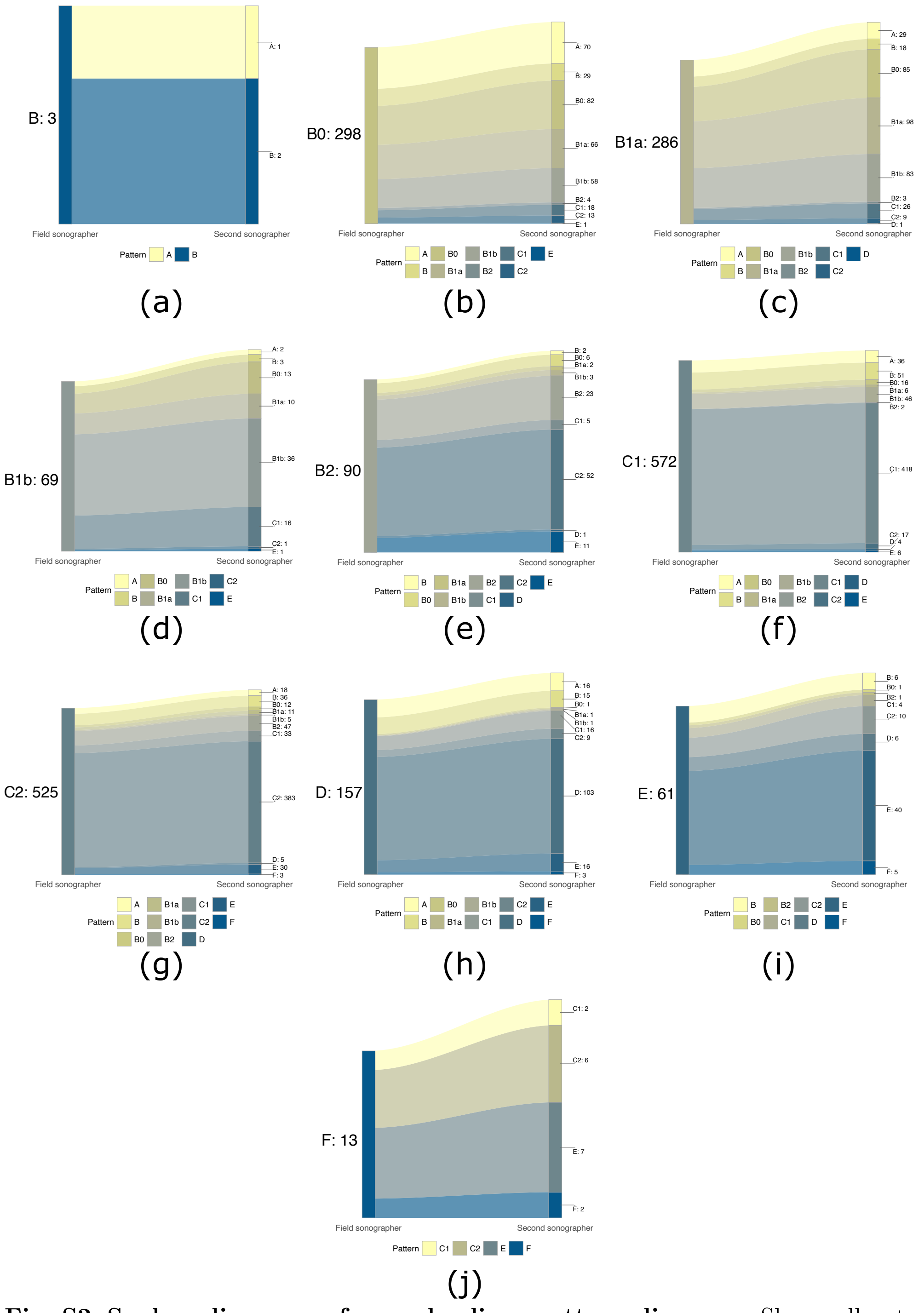


**Fig. S2**: **Sankey diagrams of granular liver pattern diagnoses** Shows all patterns assigned at second reading for each granular pattern: (a) B. (b) B0. (c) B1a. (d) B1b. (e) B2. (f) C1. (g) C2. (h) D. (i) E. (j) F.
